# Systems-level patterns in biological processes are changed under prolongevity interventions and across biological age

**DOI:** 10.1101/2022.07.11.22277435

**Authors:** Kengo Watanabe, Tomasz Wilmanski, Priyanka Baloni, Max Robinson, Gonzalo G. Garcia, Michael R. Hoopmann, Mukul K. Midha, David H. Baxter, Michal Maes, Seamus R. Morrone, Kelly M. Crebs, Charu Kapil, Ulrike Kusebauch, Jack Wiedrick, Jodi Lapidus, Jennifer C. Lovejoy, Andrew T. Magis, Christopher Lausted, Jared C. Roach, Gustavo Glusman, Steven R. Cummings, Nicholas J. Schork, Nathan D. Price, Leroy Hood, Richard A. Miller, Robert L. Moritz, Noa Rappaport

**Author notes:** Correspondence to: Leroy Hood, Noa Rappaport.

## Abstract

Aging manifests as progressive deterioration in cellular and systemic homeostasis, requiring systems-level perspectives to understand the gradual molecular dysregulation of underlying biological processes. Here, we report systems-level changes in the molecular regulation of biological processes under multiple lifespan-extending interventions in mice and across age in humans. In mouse cohorts, Differential Rank Conservation (DIRAC) analyses of liver proteomics and transcriptomics show that mechanistically distinct prolongevity interventions tighten the regulation of aging-related biological modules, including fatty acid metabolism and inflammation processes. An integrated analysis of liver transcriptomics with mouse genome-scale metabolic model supports the shifts in fatty acid metabolism. Additionally, the difference in DIRAC patterns between proteins and transcripts suggests biological modules which may be tightly regulated via cap-independent translation. In a human cohort spanning the majority of the adult lifespan, DIRAC analyses of blood proteomics and metabolomics demonstrate that regulation of biological modules does not monotonically loosen with age; instead, the regulatory patterns shift according to both chronological and biological ages. Our findings highlight the power of systems-level approaches to identifying and characterizing the biological processes involved in aging and longevity.

## Introduction

Aging manifests as progressive deterioration in cellular and systemic homeostasis. In humans, it is accompanied by an increased risk for chronic conditions such as diabetes, heart disease, neurodegeneration, and cancer^1,2^. Interventions targeting aging mechanisms could delay or moderate chronic diseases and improve health and lifespan^3^. However, aging involves diverse chemical and physiological components, posing a challenge to comprehensive understanding^4^. For instance, many studies have demonstrated key roles of nutrient-sensing pathways in aging and longevity across species, including growth hormone (GH) and insulin/insulin growth factor 1 (IGF-1), AMP-activated protein kinase (AMPK), sirtuins, and mammalian (or mechanistic) target of rapamycin (mTOR) signaling pathways^5–9^, but these nutrient-sensing pathways are intricately interconnected with each other. Given the complex and multifaceted nature of aging, systems-level approaches may provide different perspectives from single molecule-level approaches and deepen our understanding of the aging processes.

Some nutritional and pharmacological interventions consistently extend lifespan and healthspan in mouse and other animal models^3,10–12^. Nutritional interventions include calorie restriction (CR)^13^, methionine restriction (MR)^14^, and ketogenic diet^15,16^. While the number of possible “geroprotectors” has been growing^17^, pharmacological interventions whose effects on lifespan extension were robustly confirmed by the National Institute on Aging (NIA) Interventions Testing Program (ITP)^18^ include acarbose (ACA)^19–21^, canagliflozin^22^, 17α-estradiol (17aE2)^19,20,23^, glycine^24^, nordihydroguaiaretic acid^19,20,25^, Protandim® (a Nrf2 inducer)^20^, and rapamycin (Rapa)^26–28^. Rapa is the only drug found to prolong lifespan in every organism studied, including yeast, worms, flies, and mammals^29,30^. Rapa modulates nutrient-sensing pathways by inhibiting the activity of mTOR through complex formation with FK506-binding protein 12, which globally attenuates protein translation via mTOR complex 1 (mTORC1) and ultimately reduces inflammation, increases autophagy, and improves stem cell maintenance^31,32^. ACA could potentially mimic some aspects of CR^19^; it is an oral antidiabetic drug which competitively inhibits the activity of α-glucosidase enzymes to digest polysaccharides, resulting in the deceleration of sugar uptake in the gastrointestinal tract^33^. ACA treatment has been shown to extend lifespan in male mice more than in female mice^19–21^, possibly due to sex-dependent differences observed in heart, liver, and gut metabolite profiles^34,35^. 17aE2 is a stereoisomer of the dominant female sex hormone 17β-estradiol, having much weaker binding affinity to the classical estrogen receptors, stronger affinity to the brain estrogen receptor, and neuroprotective properties^36,37^. 17aE2 treatment extends lifespan in male but not in female mice^19,20,23^, potentially due to male-specific reduction of age-associated neuroinflammation^38^ and sex-specific metabolomic responses observed in liver and plasma metabolite profiles^39^. Because these prolongevity drugs were tested with standardized protocols in NIA ITP and because they have differences in primary mode of action, comparisons of their effects on molecular regulation are valuable for our understanding of aging and longevity mechanisms.

Differential Rank Conservation (DIRAC) method quantifies systemic variability in gene expression within a module (i.e., a gene set, typically defined with an a priori network or pathway) for a given set of identically treated samples (called “phenotype”)^40^. Briefly, for a given module, the DIRAC algorithm first characterizes the rank consensus of each phenotype, which is represented by binary value set for pairwise gene pairs (*i, j*) of the module to indicate whether the expression of *i*-th gene is higher than that of *j*-th gene in the phenotype-sharing samples. For each sample, the DIRAC algorithm next calculates the ratio of gene pairs whose relative ranking agrees with the rank consensus of the corresponding phenotype. Finally, the average of this ratio within the phenotype-sharing samples is a measure of how robustly the samples reflect the phenotypic gene expression pattern of the module. The module is considered “tightly” regulated within a phenotype when the samples vary little from their own consensus, because biological regulatory mechanisms or pressures must act consistently across the samples to produce such a high conservation pattern. In contrast, the module is considered “loosely” regulated within a phenotype when the samples vary considerably from their consensus, indicating a lack of conservation across the samples. For instance, a previous study applying DIRAC revealed the global loose regulation of BioCarta-defined modules in more malignant phenotypes and later stages of disease progression^40^, indicating that a loss of tight regulation characterizes the dysregulation of biological processes in cancer. Hence, the DIRAC method can be used for identifying biological modules whose regulatory patterns are changed by prolongevity interventions or aging.

In this study, we report systems-level changes in the molecular regulation of biological processes, by jointly leveraging three omics datasets of mouse cohorts including the NIA ITP-confirmed prolongevity interventions and two omics datasets of a human cohort spanning the majority of the adult lifespan (Fig. 1). We apply DIRAC analysis to mouse liver protein abundance profiles, first with predefined modules derived from Gene Ontology Biological Process (GOBP) annotations and then with unbiased modules derived from Weighted Gene Co-expression Network Analysis (WGCNA)^41,42^, and demonstrate that three lifespan-extending drugs (ACA, 17aE2, and Rapa) promoted tighter regulation of aging-related modules, such as fatty acid metabolism and inflammation processes. As a complementary approach, mouse genome-scale metabolic model (GEM)^43,44^ is developed with the three drugs-including liver transcriptomics^45^, and exhibits that multiple prolongevity interventions shifted fatty acid metabolism. In addition, comparisons of DIRAC analyses between the liver proteomics and transcriptomics suggest that biological modules were tightly regulated by the prolongevity interventions at different levels: transcription vs. post-transcription including the cap-independent translation (CIT) of specific mRNAs^46^. Finally, we explore the cross-sectional relationship between the tight module regulation and age in humans; DIRAC analyses of human plasma proteomics and metabolomics^47,48^ reveal regulatory patterns of aging-related modules according to both chronological and biological ages^48^.

**Figure 1.**
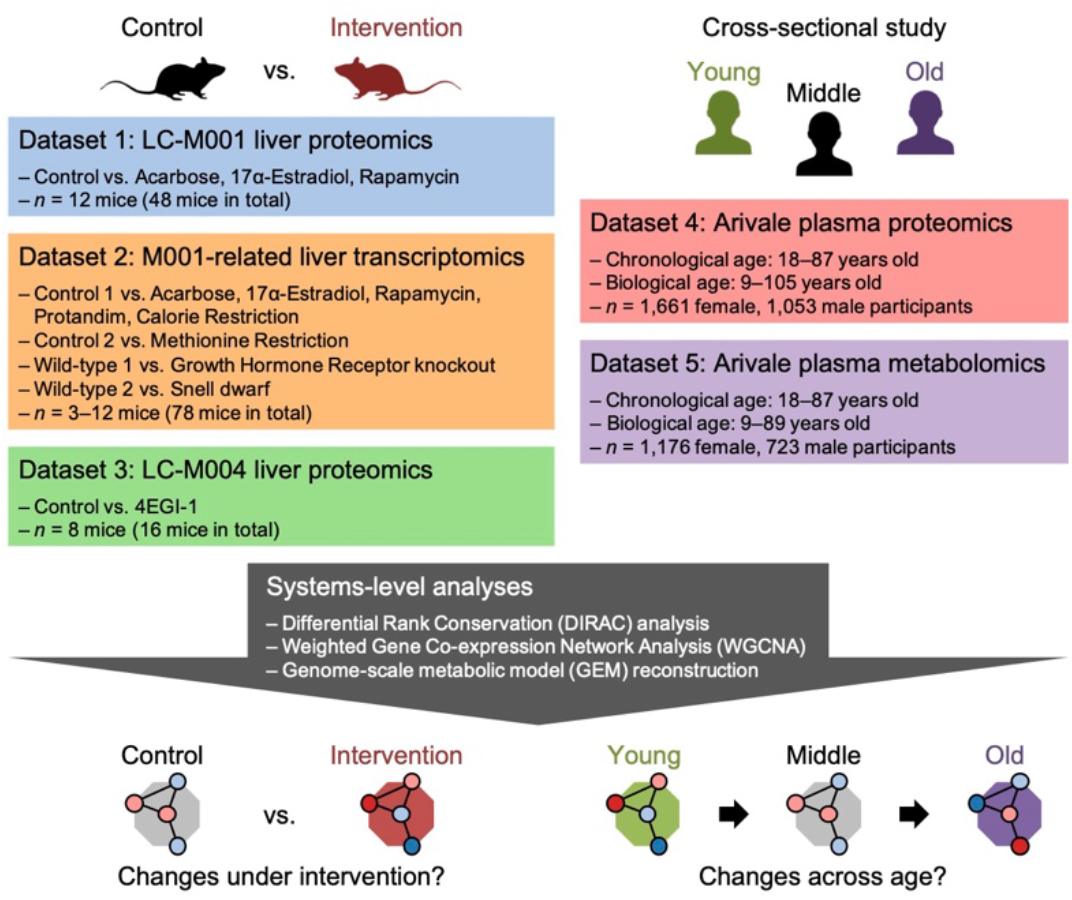
Study design overview. Schematic representation of this study. Utilizing five omics datasets and three systems-level analyses, this study addresses systems-level changes in the molecular regulation of biological processes under multiple prolongevity interventions in mice and across age in humans. LC: Longevity Consortium, a project supported by the National Institute on Aging (NIA). Dataset 2 was generated in the previous study^45^. Datasets 4 and 5 were collected through the previous studies^47,48^.

## Results

### Prolongevity interventions tightened the regulation of a priori proteomic modules

To compare the systems-level changes induced by different prolongevity interventions, we first applied DIRAC analysis to a liver proteomic dataset which was generated through a mouse prolongevity intervention experiment in the NIA Longevity Consortium (denoted “LC-M001 proteomics”; Fig. 1). In this experiment, 48 mice were either untreated (Control) or subjected to one of three lifespan-extending drug treatments (ACA, 17aE2, or Rapa), and were euthanized at 12 months (*n* = 12 (6 female, 6 male) mice per group). The design of evaluating drug effects on healthy young adult mice was motivated by the desire to reduce confounding effects of aging and of late-life diseases. In DIRAC analysis, we pooled female and male samples per intervention to calculate robust DIRAC rank consensus from small sample size, while recognizing the false negative risks for sex-dependent changes related to the sex-dependent effects of ACA and 17aE2 on lifespan extension^19–21^. For biological modules used in this DIRAC analysis, we prepared 164 a priori modules which were defined by the GOBP annotations mapped to the measured proteins (see Methods; Supplementary Data 1).

A DIRAC metric, rank conservation index (RCI)^40^, measures consistency in the relative abundance of biomolecules within a module among phenotype-sharing samples; high RCI indicates a strongly shared pattern of behavior (i.e., “tight” regulation), while low RCI indicates unpatterned behavior (i.e., “loose” regulation). ACA, 17aE2, and Rapa showed significantly higher RCI mean in the examined modules than Control (Fig. 2a), suggesting general tightening of module regulation by each of these prolongevity interventions. To identify the module changed (i.e., tightened or loosened) by any of the interventions, we assessed the intervention effect on RCI using Analysis of Variance (ANOVA) for each of the 164 modules. There were 12 significantly changed modules based on “conservatively” false discovery rate (FDR)-adjusted *P* < 0.05 (see Methods; cf. 51 modules exhibited nominal *P* < 0.05; Fig. 2b). Among these 12 changed modules, the post hoc RCI comparisons between Control and each intervention group revealed that seven, nine, and eight modules were significantly tightened by ACA, 17aE2, and Rapa, respectively (Fig. 2c), while no module was loosened. Four modules were significantly tightened under all the three interventions (Fig. 2c), which were functionally related to fatty acid β-oxidation (GO:0006635, GO:0031998) or protein-transporting to peroxisomes (GO:0016558, GO:0006625) (Fig. 2d, Supplementary Fig. 1c). Given that the primary mode of action is different between the studied drugs, this result suggests that systems-level regulation for these biological processes may be a general mechanism for lifespan extension.

**Figure 2.**
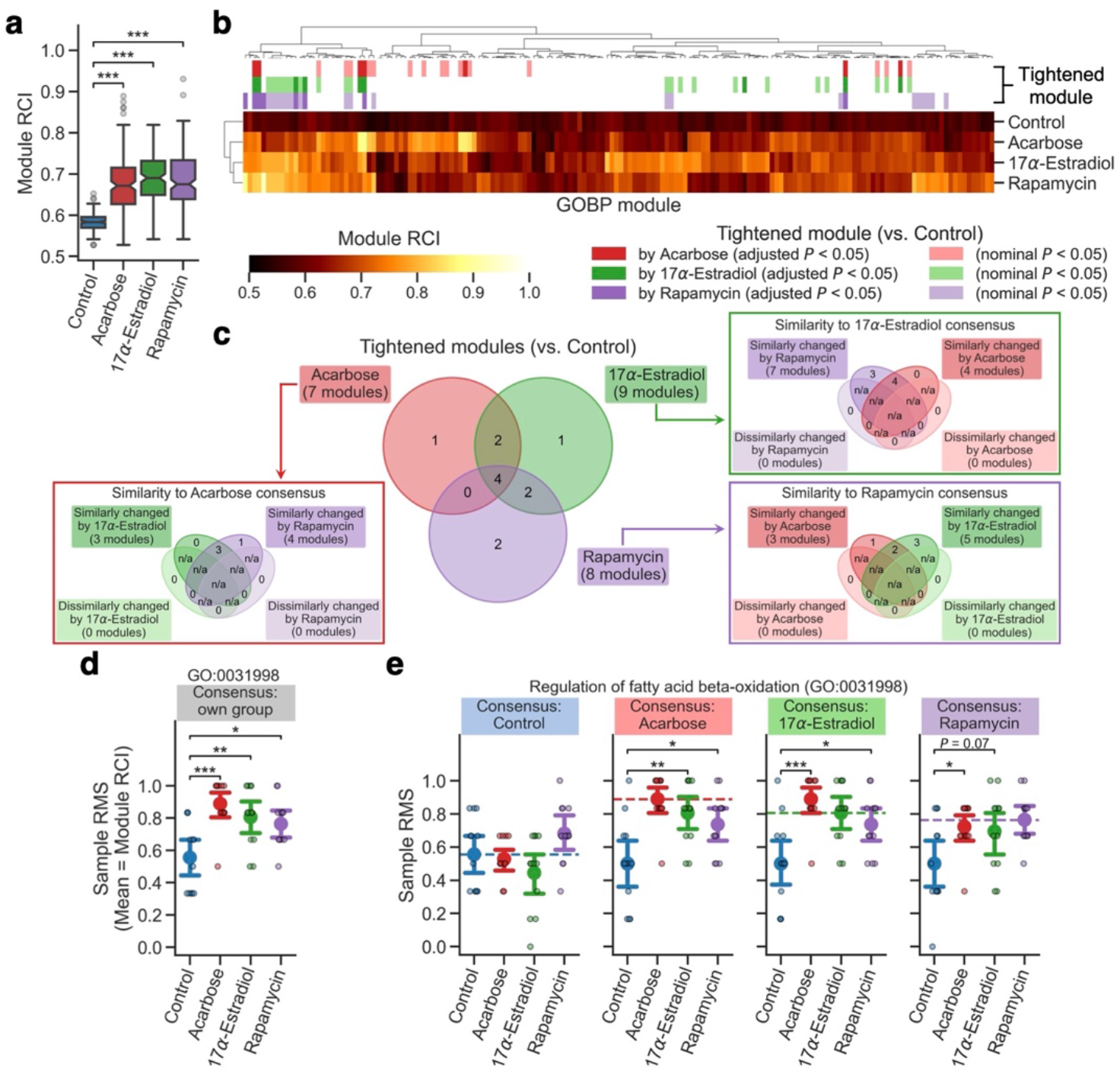
Prolongevity interventions tightened the regulation of a priori proteomic modules. **a**–**e** Differential Rank Conservation (DIRAC) analysis of the LC-M001 liver proteomics using Gene Ontology Biological Process (GOBP)-defined modules (see Supplementary Data 1 for complete results). **a, b** Overall distribution of module rank conservation index (RCI). Data (**a**): the 25^th^ percentile (*Q*_1_, box bottom), median (center line, notch: 95% confidence interval (CI) for the median), and the 75^th^ percentile (*Q*_3_, box top); whiskers span [max(*x*_min_, *Q*_1_ − 1.5 × IQR), min(*x*_max_, *Q*_3_ + 1.5 × IQR)], where *x*_min_ and *x*_max_ are the minimum and maximum, respectively, in the observed values and IQR = *Q*_3_ − *Q*_1_; *n* = 164 modules. \*\*\**P* < 0.001 by two-sided Dunnett’s test. Top color columns in **b** highlight the modules that exhibited nominal or “conservatively” false discovery rate (FDR)-adjusted *P* < 0.05 (see Methods) for the main effect of intervention on each module RCI by Analysis of Variance (ANOVA) and that exhibited significantly higher RCI in intervention group than control group (i.e., “tightened” module; *P* < 0.05 by post hoc two-sided Dunnett’s test). **c** Venn diagrams of the significantly tightened modules by each intervention (conservatively FDR-adjusted *P* < 0.05). For the tightened modules in each intervention group, sub-venn diagram indicates the modules for which the other intervention groups exhibited significantly higher or lower mean of rank matching scores (RMSs) under the rank consensus than control group (i.e., “similarly” or “dissimilarly” changed module to the consensus group, respectively; *P* < 0.05 by two-sided Dunnett’s test). n/a: logically not available. **d, e** Sample RMS distributions for an example of the tightened modules (GO:0031998, regulation of fatty acid β-oxidation). Dashed line in **e** indicates the mean of RMSs for the sample group corresponding to the rank consensus (i.e., RCI). Data: the mean (dot) with 95% CI (bar); *n* = 12 mice. \**P* < 0.05, \*\**P* < 0.01, \*\*\**P* < 0.001 by two-sided Dunnett’s test.

Although high RCI reflects a shared pattern of relative abundances within the phenotype and implies biological regulation required for the pattern, a tightly regulated module may still exhibit different relative abundance patterns under different phenotypes. Another DIRAC metric, rank matching score (RMS)^40^, allows us to compare relative abundance patterns between phenotypes, by measuring the similarity of each sample to the consensus pattern of a certain phenotype rather than measuring the consistency to the consensus pattern of the sample’s own phenotype (i.e., RCI). For instance, in *acetyl-CoA biosynthetic process from pyruvate* (GO:0006086) where higher RCI against Control was observed significantly in ACA and 17aE2 and as a tendency in Rapa (Supplementary Fig. 1a), Rapa and ACA showed significantly higher and tendentiously lower mean of RMSs, respectively, than Control under the 17aE2 rank consensus (Supplementary Fig. 1b), suggesting that Rapa changed this module similarly to 17aE2 while ACA did dissimilarly. Moreover, this Rapa’s RMS mean was comparable to 17aE2’s RMS mean under the 17aE2 rank consensus (i.e., corresponding to 17aE2’s RCI) (Supplementary Fig. 1b). These DIRAC patterns suggest two modes of tight regulation for this module: one under ACA and the other under 17aE2 and Rapa. Thus, using RMS under each group’s rank consensus, we explored the similarly tightened modules across the interventions. Among the seven, nine, and eight significantly tightened modules by ACA, 17aE2, and Rapa, three, four, and two modules were similarly changed by the other two interventions, respectively (Fig. 2c). In particular, the four consistently tightened modules across the interventions (Fig. 2c, d, Supplementary Fig. 1c) exhibited significantly higher mean of RMSs in intervention groups compared to Control under almost all the other intervention group’s rank consensus (e.g., 17aE2 and Rapa showed significantly higher RMS mean than Control under the ACA rank consensus; Fig. 2e, Supplementary Fig. 1d), suggesting that fatty acid β-oxidation and peroxisome transport were similarly tightened by mechanistically distinct prolongevity interventions and thus may be a general mechanism contributing to longevity.

Taken together, these results suggest that prolongevity interventions generally tightened the regulation of the examined proteomic modules and, in the modules related to fatty acid β-oxidation and peroxisome transport, the tightened protein expression profile was similar between different drugs.

### Prolongevity interventions tightened the regulation of data-driven proteomic modules

Given potential biases in the module definitions with GOBP terms, we inferred data-driven modules using an unsupervised clustering approach, WGCNA^41,42^. WGCNA identifies modules of highly interconnected biomolecules, relying on the overall correlation network computed from high-dimensional data. We applied WGCNA to the LC-M001 proteomics and identified nine modules, ranging in size from 66 to 839 proteins (Fig. 3a). Each WGCNA module can be characterized by the “module eigengene” (i.e., the first principal component (PC) of the protein abundance matrix for the module)^42^. To identify the module associated with any of the interventions out of the nine data-driven modules, we assessed the intervention effect on the module eigengene for each module using ANOVA model with intervention, sex, and intervention–sex interaction terms. There were no significant interaction effects in any of the nine modules, and only one module, denoted Darkgreen, exhibited a significant intervention effect (*P* = 0.00082 after the Bonferroni adjustment). The post-hoc comparison revealed that 17aE2 and Rapa, but not ACA, showed significantly higher value of the module eigengene than Control in the Darkgreen module (Fig. 3b, c), suggesting that the expression profile of the Darkgreen module was changed specifically by 17aE2 and Rapa.

**Figure 3.**
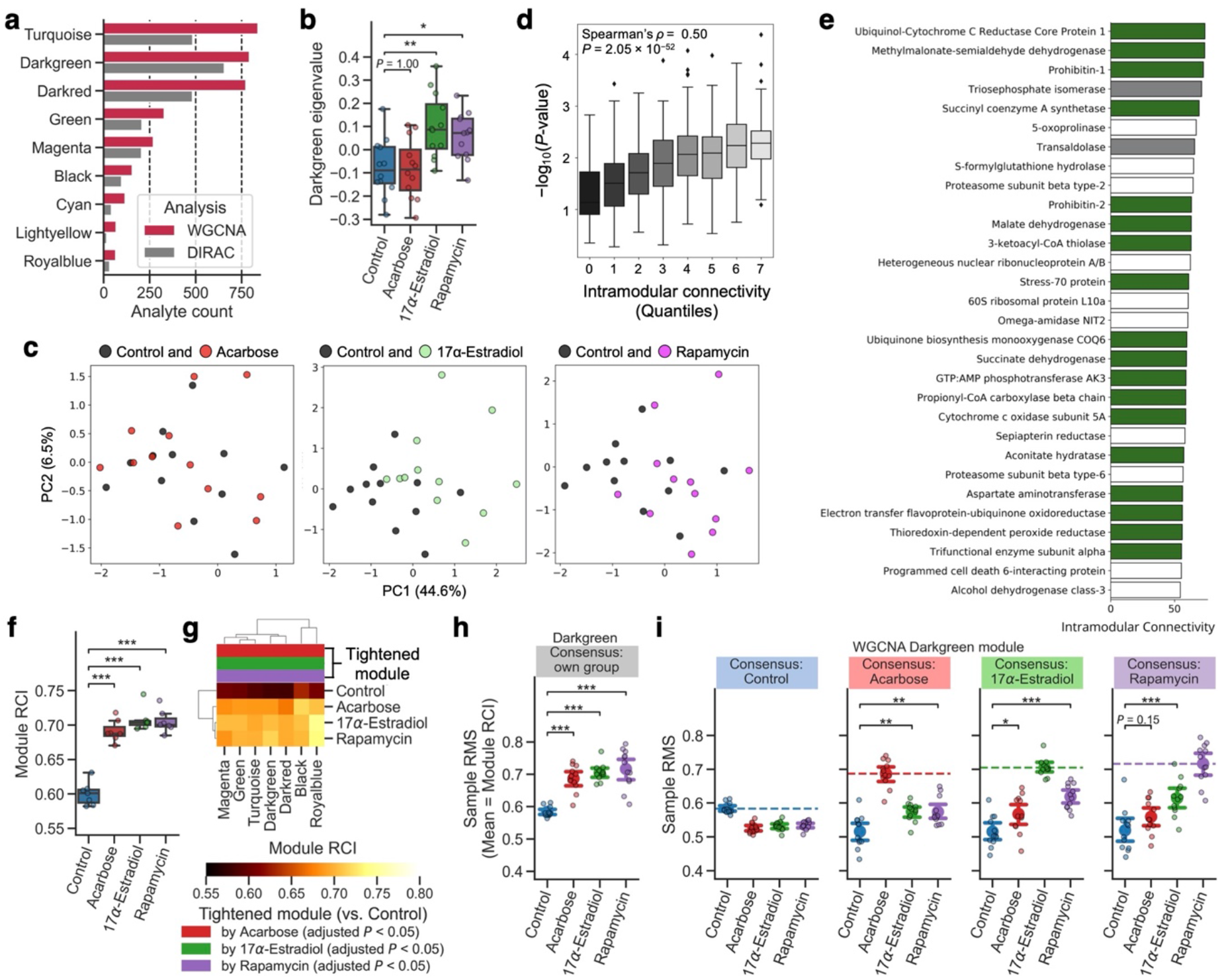
Prolongevity interventions tightened the regulation of data-driven proteomic modules. **a**–**e** Weighted Gene Co-expression Network Analysis (WGCNA) of the LC-M001 liver proteomics. **A** The number of proteins in each WGCNA-identified module. WGCNA: proteins used in WGCNA, DIRAC: proteins retained after the processing for Differential Rank Conservation (DIRAC) analysis (**f**–**i**). **b** Sample eigenvalue distributions for the Darkgreen module. Data: the 25^th^ percentile (*Q*_1_, box bottom), median (center line), and the 75^th^ percentile (*Q*_3_, box top); whiskers span [max(*x*_min_, *Q*_1_ − 1.5 × IQR), min(*x*_max_, *Q*_3_ + 1.5 × IQR)], where *x*_min_ and *x*_max_ are the minimum and maximum, respectively, in the observed values and IQR = *Q*_3_ − *Q*_1_; *n* = 12 mice. \**P* < 0.05, \*\**P* < 0.01 by two-sided Dunnett’s test. **c** Principal component (PC) analysis of each sample’s Darkgreen-module protein levels. The percentage of the axis title indicates the explained variance by the PC. **d** Relationship between the intervention effect on each protein in the Darkgreen module and their respective intramodular connectivity. The *P*-value of y-axis corresponds to the main effect of intervention on each protein level by Analysis of Variance (ANOVA). Each boxplot metric is the same with **b. e** Top 30 hub proteins within the Darkgreen module. Green, gray, and white colors correspond to mitochondrial, cytosolic metabolism-related, and other proteins, respectively. **f**–**i** DIRAC analysis of the LC-M001 liver proteomics using WGCNA-identified modules (see Supplementary Data 2 for complete results). **f, g** Overall distribution of module rank conservation index (RCI). Data (**f**): each boxplot metric is the same with **b**; *n* = 7 modules. \*\*\**P* < 0.001 by two-sided Dunnett’s test. Top color columns in **g** highlight the modules that exhibited false discovery rate (FDR)-adjusted *P* < 0.05 for the main effect of intervention on each module RCI by ANOVA and that exhibited significantly higher RCI in intervention group than control group (i.e., “tightened” module; *P* < 0.05 by post hoc two-sided Dunnett’s test). **h, i** Sample rank matching score (RMS) distributions for the Darkgreen module. Dashed line in **i** indicates the mean of RMSs for the sample group corresponding to the rank consensus (i.e., RCI). Data: the mean (dot) with 95% CI (bar); *n* = 12 mice. \**P* < 0.05, \*\**P* < 0.01, \*\*\**P* < 0.001 by two-sided Dunnett’s test.

WGCNA fits a “scale-free” network topology where the majority of nodes share relatively few edges with other nodes, while the central nodes that have high intramodular connectivity (called “hub” nodes) frequently take essential functions in the system^49^. To better understand how 17aE2 and Rapa changed the Darkgreen module structure, we assessed the relationship between the intervention effect on each protein in the Darkgreen module and their respective intramodular connectivity (see Methods). The intervention effect on each protein showed significant positive correlation with intramodular connectivity (Spearman’s *ρ* = 0.50, *P* = 2.05 × 10^−52^, Fig. 3d), suggesting that intramodular hub proteins were more strongly affected by the interventions than less connected proteins. Interestingly, 18 of the top 30 hub proteins in the Darkgreen module were mitochondrial proteins, involved in tricarboxylic acid (TCA) cycle metabolism and oxidative phosphorylation (Fig. 3e). Furthermore, prohibitin 1 (PHB1) and PHB2, 2 of the top 10 hub proteins, form the mitochondrial PHB complex, which is known to regulate fatty acid oxidation and assembly of mitochondrial respiratory complexes^50,51^, as well as to affect lifespan in *C. elegans*^52^. Collectively, our results from WGCNA revealed coordinated changes in the expression profiles of mitochondrial liver proteins that were limited to two (17aE2 and Rapa) of the three studied drugs.

We subsequently re-analyzed the LC-M001 proteomics data using the DIRAC method with seven of the nine WGCNA-identified modules (see Methods; Fig. 3a, Supplementary Data 2). ACA, 17aE2, and Rapa showed significantly higher RCI mean in the examined modules than Control (Fig. 3f), suggesting general tightening of module regulation by each of these prolongevity interventions, consistent with the initial DIRAC result based on GOBP terms (Fig. 2a). Additionally, all the seven WGCNA modules exhibited significant intervention effects on RCI in ANOVA (FDR-adjusted *P* < 0.05; Fig. 3g) and significantly higher RCI in any intervention group compared to Control in the post hoc RCI comparisons (Fig. 3g, h), suggesting that all the WGCNA modules were consistently tightened across ACA, 17aE2, and Rapa. Moreover, the Darkgreen module exhibited significantly higher mean of RMSs in intervention groups compared to Control under almost all the other intervention group’s rank consensus (e.g., 17aE2 and Rapa showed significantly higher RMS mean than Control under the ACA rank consensus; Fig. 3i), suggesting that the tightly regulated patterns are similar between the three drugs. At the same time, Rapa’s RMS mean was more similar to 17aE2’s RMS mean than to ACA’s RMS mean (e.g., 17aE2 showed higher RMS mean than ACA under the Rapa rank consensus) in the Darkgreen module (Fig. 3i), implying a difference in the tightly regulated pattern between ACA vs. 17aE2 and Rapa, in line with their effects on module expression profiles (Fig. 3b). Note that, if the tightly regulated pattern of ACA was completely different from those of 17aE2 and Rapa, ACA could have showed lower RMS mean than Control under the 17aE2 or Rapa rank consensus (cf. Supplementary Fig. 1b).

These findings suggest that the Darkgreen module was tightly regulated across all the three interventions, while also exhibiting intervention-specific effects on protein expression profiles related to mitochondrial energy metabolism. Altogether, our results indicate that the tightening of module regulation was a general signature of the prolongevity interventions within the measured proteomic space.

### Prolongevity interventions shifted the flux regulation in fatty acid metabolism

As a complementary approach to the findings from DIRAC and WGCNA analyses, we performed in silico analysis using the mouse GEM^43^ to investigate metabolic shifts associated with prolongevity interventions. GEM is a mathematical framework that leverages knowledge-base cataloging information about biochemical reactions within a system (e.g., single cell, tissue, organ), including metabolites, genes encoding catalytic enzymes, and their stoichiometry^44^. Using optimization techniques with large-scale experimental data (e.g., transcriptomics), the solved stoichiometric coefficients of each reaction allow flux prediction for metabolic reactions in the system at equilibrium^53^. Thus, GEM has been used to investigate metabolic changes in various systems and specific contexts (e.g., human cancers)^54^. Since the detected proteins in the LC-M001 proteomics did not sufficiently cover the metabolic proteins included in the mouse GEM, we utilized a mouse liver transcriptomic dataset from a previous prolongevity intervention study^45^ (referred as “M001-related transcriptomics”; Fig. 1), whose experimental design resembled the LC-M001 experiment and contained ACA, 17aE2, and Rapa treatments as prolongevity interventions. In this M001-related experiment, 78 mice were prepared for either control or one of the prolongevity interventions, including two genetically modified models (the growth hormone receptor knockout mouse (GHRKO) and the hypopituitary Snell dwarf mouse (SnellDW)), two nutritional interventions (CR and MR), and four pharmacological interventions (ACA, 17aE2, Protandim, and Rapa), and were euthanized at young adult ages depending on the intervention type (*n* = 3–12 mice per intervention group; see Methods). By integrating the M001-related transcriptomics with the mouse generic GEM^43^ for each of 78 samples, we generated 78 “context-specific” metabolic networks (i.e., GEMs constrained by each sample condition), and subsequently predicted flux values of the metabolic reactions for each context-specific GEM (see Methods). As a result, the flux values were successfully predicted for 7,930 reactions among the 10,612 reactions defined in the generic GEM (Supplementary Data 3).

To identify the reaction changed by any of the interventions, we assessed the intervention effect on the flux value using Kruskal–Wallis *H*-test for each of the 7,930 reactions. To mitigate the small sample size, we pooled samples per intervention in this analysis, while recognizing the false negative risks for age and sex-dependent changes related to the sex-dependent effects of ACA, 17aE2, and Protandim on lifespan extension^19–21^. There were 1,822 significantly changed reactions based on “conservatively” FDR-adjusted *P* < 0.05 (see Methods; cf. 2,156 reactions exhibited nominal *P* < 0.05; Fig. 4a). Among these 1,822 changed reactions, the post hoc comparisons of flux values between each intervention group and its corresponding control group revealed that 9, 1, 730, 851, and 1,015 reactions were significantly changed by ACA, Rapa, MR, GHRKO, and SnellDW, respectively (Fig. 4a–c), while no reaction was significantly changed by 17aE2, Protandim, and CR. We observed that many of these changed reactions belonged to several specific “subsystems”, analogous to the functional pathways, in the GEM system. For instance, when we focused on the central energy metabolism, 8 of the 10 changed reactions belonged to the *fatty acid oxidation* subsystem and exhibited the concordant direction of flux change across interventions (Fig. 4d). However, it was possible that the frequency of observed subsystems in the changed reactions was merely dependent on the number of mapped reactions to the subsystem, which is largely different between subsystems^43^. Hence, to interpret which subsystems in GEM were shifted by prolongevity interventions, we further performed overrepresentation analysis on the significantly changed reactions of ACA, MR, GHRKO, and SnellDW. Among the 5, 53, 60, and 57 tested subsystems that were annotated to any of the changed reactions, 0, 5, 13, and 11 subsystems were significantly enriched in the changed reactions of ACA, MR, GHRKO, and SnellDW, respectively (FDR-adjusted *P* < 0.05; Fig. 4e, Supplementary Fig. 2a, b, Supplementary Data 4). In particular, the *biotin metabolism, cholesterol metabolism, fatty acid oxidation*, and *fatty acid synthesis* subsystems were consistently enriched across MR, GHRKO, and SnellDW (Fig. 4e, Supplementary Fig. 2a, b), suggesting that these biological processes were shifted at the systems level by mechanistically distinct prolongevity interventions.

**Figure 4.**
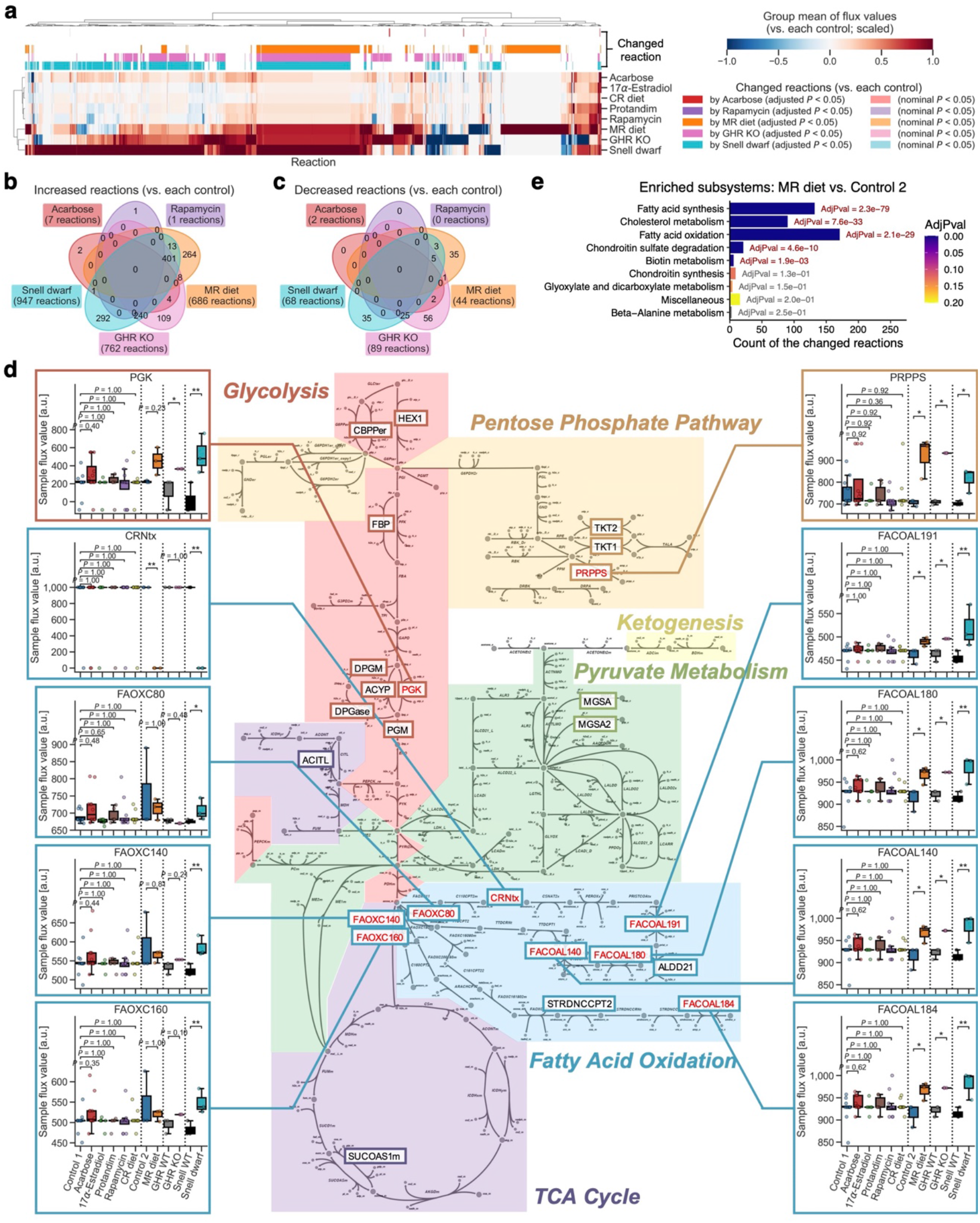
Prolongevity interventions shifted the flux regulation in fatty acid metabolism. **a**–**e** An integrated analysis of the M001-related liver transcriptomics^45^ with mouse genome-scale metabolic model (GEM; see Supplementary Data 3 and 4 for complete results). CR: calorie restriction; MR: methionine restriction; GHR: growth hormone receptor; WT: wild-type; KO: knockout; Control 1: control for Acarbose, 17α-Estradiol, Protandim, Rapamycin, and CR diet; Control 2: control for MR diet. **a** Change in the group mean of flux values for each reaction. The presented group mean value was centered and scaled (see Methods); i.e., its positive value corresponds to an increase in the mean of flux values compared to corresponding control group, and vice versa. Among 7,930 reactions whose flux values were successfully predicted across all samples (78 mice), presented are the 2,156 reactions that exhibited nominal or “conservatively” false discovery rate (FDR)-adjusted *P* < 0.05 (see Methods) for the main effect of intervention on each flux median by Kruskal–Wallis *H*-test and that exhibited significantly different flux median in intervention group from control group (i.e., “changed” reaction; *P* < 0.05 by post hoc two-sided Dunn’s test with the Holm–Bonferroni adjustment). These significantly changed modules are highlighted in the top color columns per intervention group, except for 17α-Estradiol, Protandim, and CR diet due to no significantly changed reactions by them. **b, c** Venn diagrams of the significantly changed reactions by each intervention (conservatively FDR-adjusted *P* < 0.05). **d** Changed reactions within the central energy metabolism. The 25 reaction IDs highlighted in the diagram are the reactions that had the predicted flux values across all samples. Flux value distributions are presented for the 10 reactions that exhibited nominal *P* < 0.05 by the aforementioned Kruskal–Wallis *H*-test. Data: the 25^th^ percentile (*Q*_1_, box bottom), median (center line), and the 75^th^ percentile (*Q*_3_, box top); whiskers span [max(*x*_min_, *Q*_1_ − 1.5 × IQR), min(*x*_max_, *Q*_3_ + 1.5 × IQR)], where *x*_min_ and *x*_max_ are the minimum and maximum, respectively, in the observed values and IQR = *Q*_3_ − *Q*_1_; *n* = 12 (Control 1, Acarbose, Rapamycin, CR diet), 6 (17α-Estradiol, Protandim), 3 (the others) mice. \**P* < 0.05, \*\**P* < 0.01 by two-sided Dunn’s test with the Holm–Bonferroni adjustment. **e** Shifted subsystems by MR diet. Significance of the shifted subsystems was assessed using enrichment analysis on the significantly changed reactions (conservatively FDR-adjusted *P* < 0.05) while adjusting multiple hypotheses with the Benjamini–Hochberg method. Only the subsystems that exhibited nominal *P* < 0.05 are presented. AdjPval: FDR-adjusted *P*-value from the enrichment analysis.

Given that MR has been often discussed as if it was a form of CR, we addressed the difference in reaction flux between CR and MR. Among the 1,822 reactions changed by any of the interventions, the post hoc comparisons of flux values among CR, MR, and their corresponding control groups revealed that 0, 1,081, and 1,229 reactions were significantly different in CR vs. control, MR vs. control, and MR vs. CR, respectively (Supplementary Fig. 2c–f). Overrepresentation analysis revealed that 11 subsystems were significantly enriched in the different reactions between CR and MR (FDR-adjusted *P* < 0.05; Supplementary Fig. 2g), implying that MR shifted these subsystems at the systems level in a different manner from CR.

In summary, our in silico analysis differentiated the metabolic effects of different prolongevity interventions, and implied that multiple prolongevity interventions concordantly shifted fatty acid metabolism at the systems level.

### Prolongevity interventions likely tightened the module regulation partly through cap-independent translation

To further investigate the tightening effects of prolongevity interventions on proteomic modules (Fig. 2, 3), we applied DIRAC analysis to the M001-related transcriptomics^45^. Again, we pooled samples per intervention to calculate robust DIRAC rank consensus, and analyzed only three prolongevity interventions (ACA, Rapa, and CR) and their corresponding control (Control) based on sample size (*n* = 12 (6 female, 6 male) mice per group). Using 3,747 a priori modules defined by the GOBP annotations mapped to the measured transcripts (see Methods; Supplementary Data 5), we found that ACA, Rapa, and CR showed significantly higher RCI mean in the examined modules than Control (Fig. 5a). This result suggests the general tightening of module regulation within the measured transcriptomic space, as well as within the measured proteomic space (Fig. 2a, 3f). We next assessed the intervention effect on RCI using ANOVA for each of the 3,747 modules, and identified 1,829 significantly changed modules by any of the interventions based on “conservatively” FDR-adjusted *P* < 0.05 (see Methods; cf. 2,107 modules exhibited nominal *P* < 0.05; Supplementary Fig. 3a). Among these 1,829 changed modules, the post hoc RCI comparisons between Control and each intervention group revealed that 828, 432, and 1,789 modules were significantly tightened by ACA, Rapa, and CR, respectively (Supplementary Fig. 3a), while no module was loosened. Subsequently, using RMS under each group’s rank consensus, we explored the similarly tightened modules across the interventions. Among the 828, 432, and 1,789 significantly tightened modules by ACA, Rapa, and CR, 35, 19, and 12 modules were similarly changed by the other two interventions, respectively (Supplementary Fig. 3b). For instance, *ubiquitin-dependent protein catabolic process* (GO:0006511), a consistently tightened module across the interventions (Supplementary Fig. 3c), exhibited significantly higher mean of RMSs in intervention groups compared to Control under almost all the other intervention group’s rank consensus (e.g., Rapa and CR showed significantly higher RMS mean than Control under the ACA rank consensus; Supplementary Fig. 3d), suggesting that this module was similarly tightened in transcripts by mechanistically distinct prolongevity interventions.

**Figure 5.**
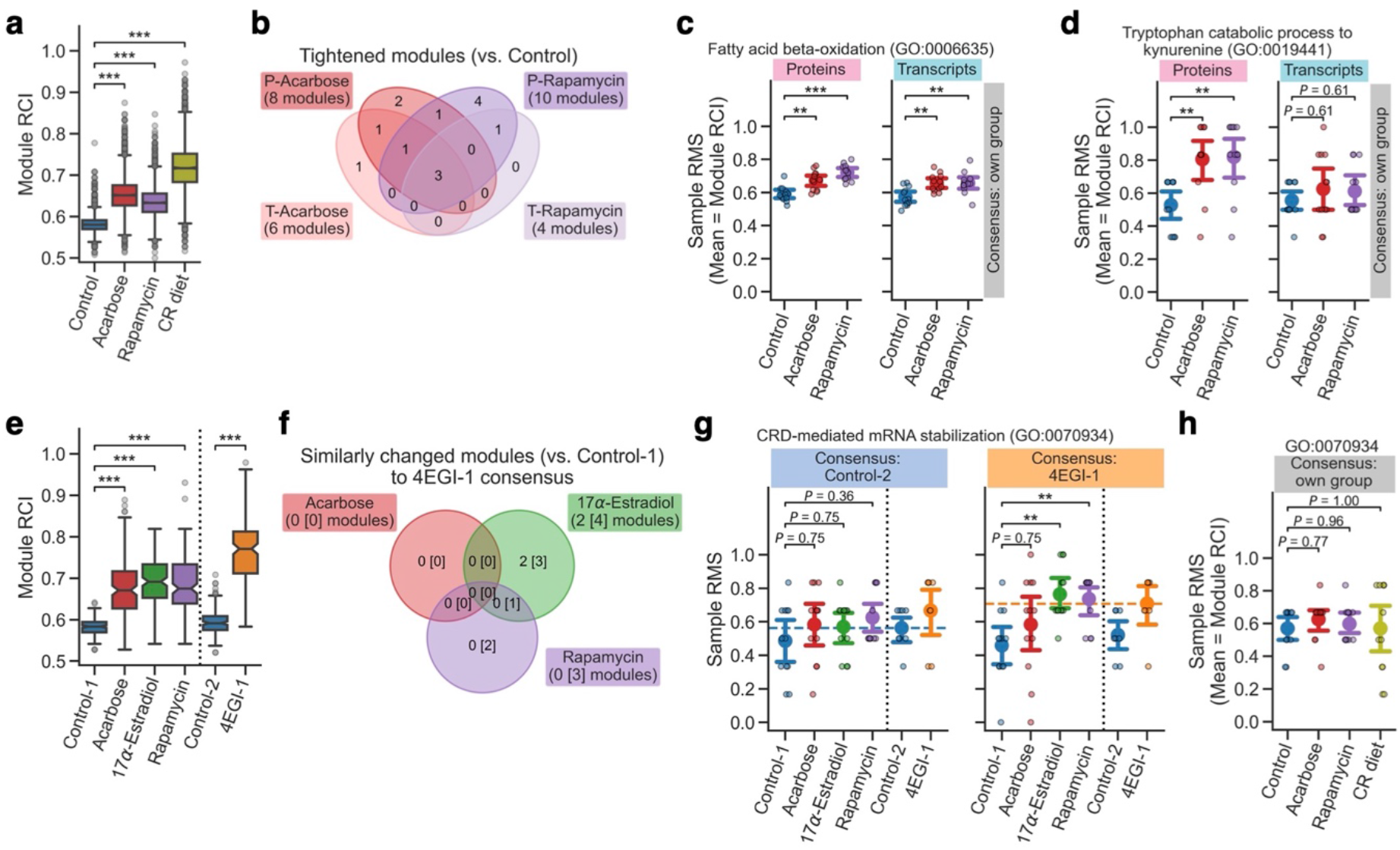
Prolongevity interventions likely tightened the module regulation partly through cap-independent translation. **a** Differential Rank Conservation (DIRAC) analysis of the M001-related liver transcriptomics using Gene Ontology Biological Process (GOBP)-defined modules (see Supplementary Data 5 for complete results). Presented is overall distribution of module rank conservation index (RCI). CR: calorie restriction. Data: the 25^th^ percentile (*Q*_1_, box bottom), median (center line, notch: 95% confidence interval (CI) for the median), and the 75^th^ percentile (*Q*_3_, box top); whiskers span [max(*x*_min_, *Q*_1_ − 1.5 × IQR), min(*x*_max_, *Q*_3_ + 1.5 × IQR)], where *x*_min_ and *x*_max_ are the minimum and maximum, respectively, in the observed values and IQR = *Q*_3_ − *Q*_1_; *n* = 3,747 modules. \*\*\**P* < 0.001 by two-sided Dunnett’s test. **b**–**d** Comparison of DIRAC results between the LC-M001 liver proteomics and the M001-related liver transcriptomics (see Supplementary Data 6 for complete results). **b** Venn diagram of the modules that exhibited “conservatively” false discovery rate (FDR)-adjusted *P* < 0.05 (see Methods) for the main effect of intervention on each module RCI by Analysis of Variance (ANOVA) and that exhibited significantly higher RCI in intervention group than control group (i.e., “tightened” module; *P* < 0.05 by post hoc two-sided Student’s *t*-tests with the Holm–Bonferroni adjustment). P: proteomics, T: transcriptomics. **c, d** Sample rank matching score (RMS) distributions for an example of the tightened modules in both proteins and transcripts (**c**; GO:0006635, fatty acid β-oxidation) or the tightened modules only in proteins (**d**; GO:0019441, tryptophan catabolic process to kynurenine). Data: the mean (dot) with 95% CI (bar); *n* = 12 mice. \*\**P* < 0.01, \*\*\**P* < 0.001 by two-sided Student’s *t*-tests with the Holm–Bonferroni adjustment. **e**–**g** DIRAC analysis of the LC-M001 and LC-M004 liver proteomics using GOBP-defined modules (see Supplementary Data 7 for complete results). Control 1: control for Acarbose, 17α-Estradiol, and Rapamycin; Control 2: control for 4EGI-1. **e** Overall distribution of module RCI. Data: each boxplot metric is the same with **a**; *n* = 153 modules. \*\*\**P* < 0.001 by two-sided Student’s *t*-tests with the Holm–Bonferroni adjustment. **f** Venn diagram of the modules that exhibited nominal or “conservatively” FDR-adjusted *P* < 0.05 (see Methods) for the main effect of intervention on each module mean of RMSs under 4EGI-1 rank consensus by ANOVA and that exhibited significantly higher mean of RMSs in intervention group than control group (i.e., “similarly” changed module to the 4EGI-1 group; *P* < 0.05 by post hoc two-sided Student’s *t*-tests with the Holm–Bonferroni adjustment). The number in square brackets corresponds to the similarly changed modules (nominal *P*-value < 0.05). **g, h** Sample RMS distributions for an example of the similarly tightened modules (GO:0070934, coding region instability determinant (CRD)-mediated mRNA stabilization) in proteins (**g**) or transcripts (**h**). Dashed line in **g** indicates the mean of RMSs for the sample group corresponding to the rank consensus (i.e., RCI). Data: the mean (dot) with 95% CI (bar); *n* = 8 (Control-2, 4EGI-1), 12 (the others) mice. \*\**P* < 0.01 by two-sided Student’s *t*-tests with the Holm–Bonferroni adjustment (**g**) or Dunnett’s test (**h**).

To directly compare the DIRAC results between the LC-M001 proteomics and the M001-related transcriptomics, we focused on the two interventions (ACA and Rapa) and the 147 GOBP modules that were used in both omics results (Supplementary Data 6), and re-assessed the intervention effect on RCI using ANOVA for each of the 147 modules and each omics. There were 10 and 5 significantly changed modules by any of the interventions in proteins and transcripts, respectively, based on “conservatively” FDR-adjusted *P* < 0.05 (see Methods). Among these changed modules, the post hoc RCI comparisons between Control and each intervention group revealed that 8, 6, 10, and 4 modules were significantly tightened by ACA in proteins, ACA in transcripts, Rapa in proteins, and Rapa in transcripts, respectively (Fig. 5b). Interestingly, the modules that were significantly tightened by ACA and Rapa in both proteins and transcripts were three modules related to fatty acid β-oxidation (GO:0006635), retrograde transport (GO:1990126), or interleukin 7 (GO:0098761) (Fig. 5b, c). This result suggests that these modules were tightened by the prolongevity interventions via transcription-level changes with concordant changes of proteomic profiles. At the same time, we also observed seven modules which were tightened specifically in proteins (Fig. 5b). In particular, *tryptophan catabolic process to kynurenine* (GO:0019441) exhibited significantly higher RCI across interventions compared to Control specifically in proteins (Fig. 5d), suggesting that this module was tightened by ACA and Rapa in the proteomic profile but not in the transcriptomic profile. This inconsistency may reflect post-transcriptional regulatory mechanisms that can affect protein profiles beyond transcriptional changes. For instance, since the abundance of a protein is determined by both its synthesis and degradation rates, a difference in “proteostasis”, whose loss is known as an aging signature^1,55^, can lead to the change in protein abundance without a change in transcript abundance.

Likewise, CIT^46^ can be a possible post-transcriptional mechanism to explain the inconsistency between proteins and transcripts. In contrast to the standard cap-dependent translation, CIT does not require the interaction of the eukaryotic initiation factor 4E (eIF4E) complex with 5′ cap of mRNA; *N*^6^-methyladenosine (m^6^A) modification in 5′ untranslated regions of mRNA can trigger the recruitment of specific initiation and elongation factors, followed by the selective translation of m^6^A-tagged mRNAs. Previous studies have shown the upregulated translation of a subset of mRNAs via CIT in long-lived endocrine mutant mice^56^ and similar increases of CIT in mice treated with ACA, 17aE2, or Rapa^57^. We therefore tested if CIT could explain the difference in module regulation between proteins and transcripts, by jointly applying DIRAC analysis to the LC-M001 proteomics and another liver proteomic dataset which was generated through a mouse CIT experiment (denoted “LC-M004 proteomics”; Fig. 1). In this experiment, 16 mice were treated with either solvent (Control-2) or 4EGI-1, a synthetic small compound which inhibits the eIF4E–eIF4G interaction and thereby blocks cap-dependent translation and enhances CIT^58^, and were euthanized at young adult ages (*n* = 8 (4 female, 4 male) mice per group). To directly compare the DIRAC results across the LC-M001 and LC-M004 proteomics, we focused on 153 GOBP modules for this analysis, which were mapped to the measured proteins in both datasets (see Methods; Supplementary Data 7). Consistent with the elevated module tightness in ACA, 17aE2, and Rapa against their corresponding control (Control-1), 4EGI-1 showed significantly higher RCI mean in the examined modules compared to Control-2 (Fig. 5e), implying general tightening of module regulation by the CIT enhancement. To reveal the similarity of module regulation between prolongevity interventions and 4EGI-1, we calculated RMSs under the rank consensus of Control-2 and 4EGI-1 for the LC-M001 groups (Control-1, ACA, 17aE2, and Rapa), and assessed the intervention effect on the RMS mean using ANOVA for each of the 153 modules and each rank consensus. There were four and seven significantly changed modules by any of the interventions under the Control-2 and 4EGI-1 rank consensus, respectively, based on “conservatively” FDR-adjusted *P* < 0.05 (see Methods; cf. 24 and 41 modules exhibited nominal *P* < 0.05, respectively). Among these seven changed modules under the 4EGI-1 rank consensus, the post hoc comparisons for the RMS mean between Control-1 and each intervention group revealed that two, one, and three modules were changed “dissimilarly” to the 4EGI-1 consensus by ACA, 17aE2, and Rapa, respectively (Supplementary Fig. 3e). For instance, in *RIG-I signaling pathway* (GO:0039529), all the ACA, 17aE2, and Rapa showed significantly lower mean of RMSs than Control-1 under the 4EGI-1 consensus (Supplementary Fig. 3f). Given that this module was similarly tightened across the interventions in proteins (Supplementary Fig. 1e, f) while ACA and Rapa did not show the significant RCI difference from control in transcripts (Supplementary Fig. 3g), this result suggests that *RIG-I signaling pathway* may be tightened in proteins via post-transcriptional regulation other than CIT. In contrast, the post hoc RMS mean comparisons for the seven changed modules also revealed that two modules were changed “similarly” to the 4EGI-1 consensus by 17aE2 (Fig. 5f); e.g., *mitochondrial ATP synthesis coupled proton transport* (GO:0042776) exhibited significantly higher mean of RMSs in 17aE2 compared to Control-1 (Supplementary Fig. 3h). Because the regulatory pattern of 17aE2 in transcripts was not available and because our *P*-value adjustment for multiple hypotheses was conservative (see Methods), we also checked the 41 changed modules based on nominal *P* < 0.05 under the 4EGI-1 rank consensus. The post hoc RMS mean comparisons for these 41 changed modules revealed that four and three modules were changed “similarly” to the 4EGI-1 consensus by 17aE2 and Rapa, respectively (Fig. 5f). Remarkably, in *coding region instability determinant (CRD)- mediated mRNA stabilization* (GO:0070934) and *positive regulation of RNA polymerase II transcription preinitiation complex (PIC) assembly* (GO:0045899), Rapa showed significantly higher mean of RMSs than Control-1 under the 4EGI-1 rank consensus (Fig. 5g, Supplementary Fig. 3i), while Rapa did not show the significant RCI difference from control in transcripts (Fig. 5h, Supplementary Fig. 3j), suggesting that regulation of these processes was modified by Rapa likely via CIT.

Altogether, these findings suggest that the tightening of module regulation was a general signature of the prolongevity interventions even within the measured transcriptomic space and that the tightened modules in proteins were achieved through both transcriptional and post-transcriptional regulation, potentially including augmented CIT.

### Module regulation was changed across chronological and biological ages

Our results observed in mice demonstrate that the molecular regulation of biological processes is modifiable at the systems level. To address the systems-level dynamics through lifetime in humans, we investigated the cross-sectional relationship between the tight module regulation and age by applying DIRAC analysis to a plasma proteomic dataset which was collected through the Arivale program^47,48^ (Fig. 1). This cohort consisted of community-dwelling adults ranging from 18 to 89 years old, who were not screened for any particular disease, and we stratified this cohort into deciles per sex by chronological age (referred as “CA10”; Supplementary Fig. 4a). Setting these CA10 groups as the group unit for the rank consensus, we calculated DIRAC metrics for 19 a priori modules which were defined by the GOBP annotations mapped to the measured proteins (see Methods; Supplementary Data 8). The RCI median in the examined modules gradually decreased along an aging gradient in younger groups, while this overall trend was reversed in older groups (Fig. 6a). We randomized the sample–group correspondence to calculate an empirical null-hypothesis distribution, and confirmed that the RCI median was significantly higher than expected in almost all the CA10 groups (Supplementary Fig. 4b), suggesting that the examined modules were generally under the tight regulation especially in the youngest group (Q1) and the oldest group (Q10). Next, we compared the similarity of module regulation across chronological age using RMS under the Q1 or Q10 rank consensus. The module RMS mean showed significant negative and positive correlations with the quantile order of CA10 groups under the Q1 and Q10 rank consensus, respectively (Spearman’s *ρ* = −0.78 (Q1, female), −0.40 (Q1, male), 0.70 (Q10, female), 0.60 (Q10, male); Fig. 6b), suggesting that module regulation was generally more similar between closer CA10 groups and the regulatory patterns were vastly dissimilar between Q1 and Q10. To identify the module whose regulation similarity to Q1 or Q10 was associated with chronological age, we regressed the RMS under the Q1 or Q10 consensus to chronological age with Body Mass Index (BMI) and ancestry PCs as covariates for each of the 19 modules, each rank consensus, and each sex. There were 18 and 13 modules exhibiting a significant negative association under the Q1 rank consensus and 17 and 18 modules exhibiting a significant positive association under the Q10 rank consensus for female and male, respectively, based on “conservatively” FDR-adjusted *P* < 0.05 (see Methods; Fig. 6c). For instance, *neutrophil chemotaxis* (GO:0030593) exhibited significant negative and positive associations of the RMS with chronological age in both sexes under the Q1 and Q10 rank consensus, respectively (Fig. 6d), suggesting that this module was gradually changed across chronological age and its regulatory pattern was vastly dissimilar between younger and older individuals. These results suggest that the general tightness of module regulation decreased with chronological age up until midlife but then increased during older stage and that the tight patterns of module regulation were different between younger and older individuals.

**Figure 6.**
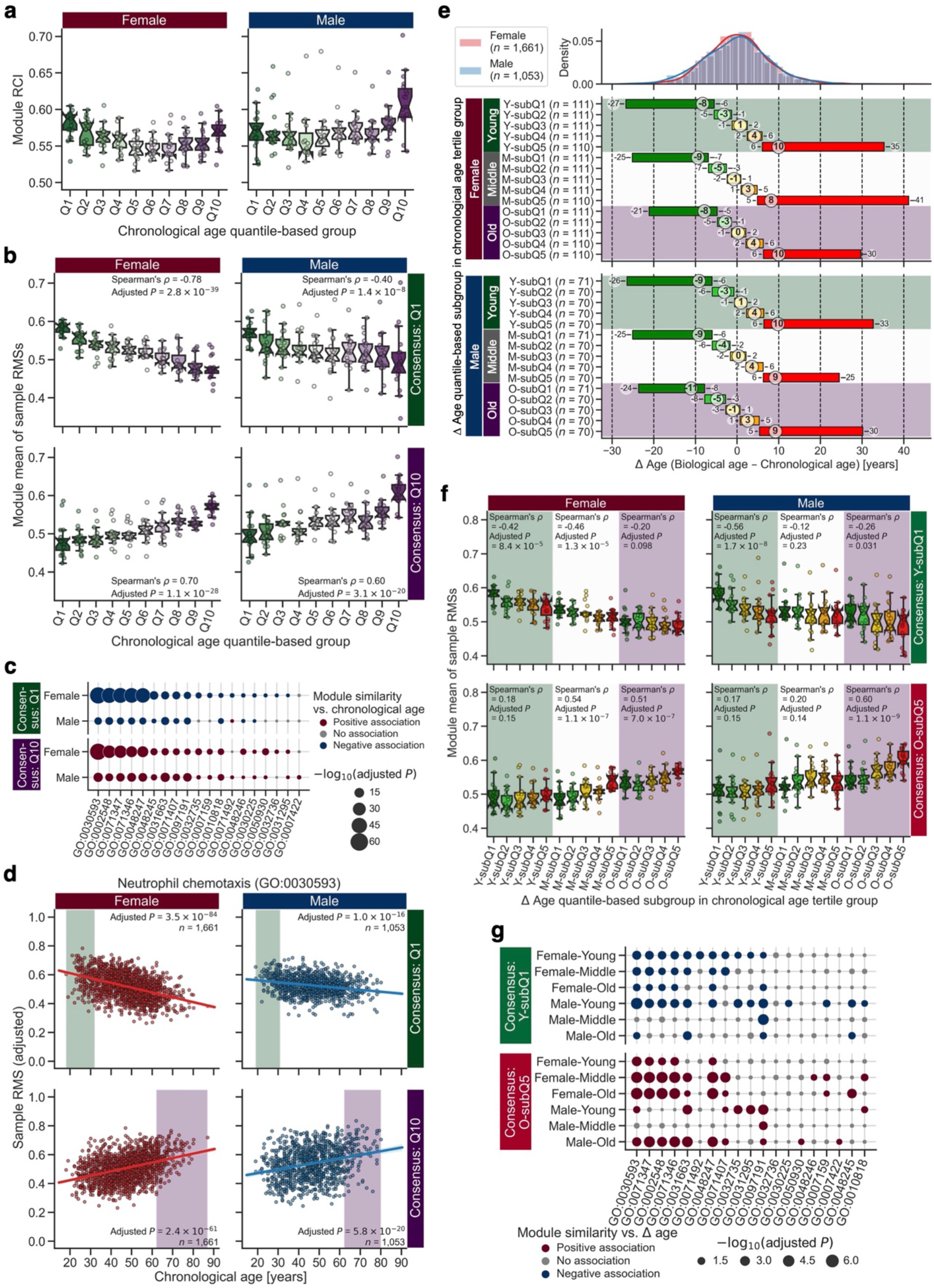
Module regulation was changed across chronological and biological ages. **a**–**g** Differential Rank Conservation (DIRAC) analysis of the Arivale plasma proteomics, with the stratified groups by chronological age (**a**–**d**) or Δ age (**e**–**g**), using Gene Ontology Biological Process (GOBP)-defined modules (see Supplementary Data 8 for complete results). **a** Overall distribution of module rank conservation index (RCI). Data: the 25^th^ percentile (*Q*_1_, box bottom), median (center line, notch: 95% confidence interval (CI) for the median), and the 75^th^ percentile (*Q*_3_, box top); whiskers span [max(*x*_min_, *Q*_1_ − 1.5 × IQR), min(*x*_max_, *Q*_3_ + 1.5 × IQR)], where *x*_min_ and *x*_max_ are the minimum and maximum, respectively, in the observed values and IQR = *Q*_3_ − *Q*_1_; *n* = 19 modules. **b** Overall distributions of module mean of rank matching scores (RMSs) under the rank consensus of the youngest (Q1) or oldest (Q10) group. Data: each boxplot metric is the same with **a**; *n* = 19 modules. *P*-value for Spearman’s correlation was adjusted across sexes with the Holm–Bonferroni method. **c** Association between module similarity to the Q1 or Q10 group and chronological age. For each module, each rank consensus, and each sex, significance of the association was assessed using ordinary least squares (OLS) linear regression with Body Mass Index (BMI) and ancestry principal components (PCs) as covariates while “conservatively” adjusting multiple hypotheses with the Benjamini–Hochberg method (see Methods). **d** An example of the significant association in **c** (GO:0030593, neutrophil chemotaxis). In each subplot, the adjusted sample RMS with the covariates (i.e., mean ± residual) is plotted, and the colored line and background correspond to the OLS linear regression line with 95% CI and the range of rank consensus group, respectively. **e** Distribution of Δ age in the Δ age-stratified groups (see Supplementary Fig. 4c for the chronological age distribution). The numbers in the subplots indicate the minimum–median–maximum of each group. **f** Overall distributions of module mean of RMSs under the rank consensus of the chronologically and biologically youngest (Y-subQ1) or oldest (O-subQ5) group. Data: each boxplot metric is the same with **a**; *n* = 19 modules. Spearman’s correlation was assessed for each of chronological age tertile (CA3) groups, and its *P*-value was adjusted across CA3 groups and sexes with the Holm–Bonferroni method. **g** Association between module similarity to the Y-subQ1 or O-subQ5 group and Δ age. For each module, each rank consensus, each sex, and each CA3 group, significance of the association was assessed using OLS linear regression with chronological age, BMI, and ancestry PCs as covariates while “conservatively” adjusting multiple hypotheses with the Benjamini–Hochberg method (see Methods).

To validate these findings in another dimensional space, we performed DIRAC analysis on a plasma metabolomic dataset of the Arivale cohort (Fig. 1). Since the dataset availability was different between participants, we re-defined CA10 groups for this analysis (Supplementary Fig. 5a). Because functional annotations to define metabolite modules are limited, we used the nine data-driven metabolomic modules identified by WGCNA (Supplementary Fig. 5b, Supplementary Data 9). Again, the RCI median in the examined modules exhibited the “U-shaped” transition with respect to CA10 group (Supplementary Fig. 5c), and the module RMS mean showed significant negative and positive correlations with the quantile order of CA10 groups under the Q1 and Q10 rank consensus, respectively (Spearman’s *ρ* = −0.90 (Q1, female), −0.76 (Q1, male), 0.87 (Q10, female), 0.66 (Q10, male); Supplementary Fig. 5d). Subsequently, we regressed the RMS under the Q1 or Q10 consensus to chronological age with BMI and ancestry PCs as covariates for each of the nine modules, each rank consensus, and each sex. All the nine modules exhibited significant negative and positive associations under the Q1 and Q10 rank consensus for both sexes, respectively, based on FDR-adjusted *P* < 0.05 (Supplementary Fig. 5e, f). Therefore, in line with the examined proteomic space, the similar associations between the tightness of module regulation and chronological age were observed in the examined metabolomics space.

Previously, a multiomic estimate for chronological age (biological age) has been calculated for the Arivale cohort^48^. Importantly, the difference between chronological and biological ages (Δ age) was a more accurate metric of wellness than chronological age (i.e., negative and positive Δ ages indicated healthier and unhealthier conditions than chronologically expected, respectively) and modifiable (i.e., lifestyle intervention decreased Δ age). Hence, we further explored the relationships between module regulation and health conditions by re-performing DIRAC analysis on the Arivale proteomics with Δ age-stratified groups; we divided the Arivale cohort into tertiles per sex by chronological age (referred as “CA3”) and further stratified each CA3 group into five subgroups by Δ age (referred as “DA5”; Fig. 6e, Supplementary Fig. 4c). Under the rank consensus of the most negative Δ age subgroup in young CA3 group (Y-subQ1) and the most positive Δ age subgroup in old CA3 group (O-subQ5), the module RMS mean showed significant negative and positive correlations with the quantile order of DA5 subgroups in young and old CA3 groups, respectively (Spearman’s *ρ* = −0.42 (Y-subQ1, young female), −0.56 (Y-subQ1, young male), 0.51 (O-subQ5, old female), 0.60 (O-subQ5, old male); Fig. 6f). Moreover, these negative and positive correlations between the module RMS mean and the quantile order of DA5 subgroups under the Y-subQ1 and O-subQ5 rank consensus, respectively, were observed in the other CA3 groups as a tendency or with statistical significance based on the Holm–Bonferroni adjustment (Fig. 6f). These results suggest that module regulation was generally more similar between closer DA5 groups and the regulatory patterns were vastly dissimilar between Y-subQ1 and O-subQ5. To identify the module whose regulation similarity to Y-subQ1 or O-subQ5 was associated with Δ age, we regressed the RMS under the Y-subQ1 or O-subQ5 consensus to Δ age with chronological age, BMI, and ancestry PCs as covariates for each of the 19 modules, each rank consensus, each sex, and each CA3 group. There were 11, 7, 6, 13, 1, and 4 modules exhibiting a significant negative association under the Y-subQ1 rank consensus and 5, 10, 9, 7, 1, and 10 modules exhibiting a significant positive association under the O-subQ5 rank consensus for female young, female middle, female old, male young, male middle, and male old CA3 groups, respectively, based on “conservatively” FDR-adjusted *P* < 0.05 (see Methods; Fig. 6g). For instance, *neutrophil chemotaxis* (GO:0030593) exhibited significant negative and positive associations of the RMS with Δ age in both sexes and all the CA3 groups, except for male middle CA3 group, under the Y-subQ1 and O-subQ5 rank consensus, respectively (Supplementary Fig. 4d), suggesting that this module was gradually changed across Δ age and its regulatory pattern was vastly dissimilar between biologically younger and biologically older individuals. Likewise, we re-performed DIRAC analysis on the Arivale metabolomics with Δ age-stratified groups, and observed the same association patterns between the RMS under the Y-subQ or O-subQ5 rank consensus and Δ age (Supplementary Fig. 6).

Altogether, these results imply that the regulatory patterns of proteomic and metabolomic modules shifted depending on both chronological and biological ages and that the tight module regulation representatively corresponded to a healthier state in the young stage but an unhealthier state in the old stage.

## Discussion

Studies in invertebrate organisms and mice have shown multiple ways to extend lifespan and postpone age-related diseases^3,10–12^. Aging can be slowed, and healthspan can be extended, by mutation of individual genes, dietary restrictions, or oral administration of compounds. Data are becoming available to determine which of the many cellular and molecular traits modified by each of these interventions are shared across slow-aging models and which are less universal. Elucidation of the physiological and cellular mechanisms of effective interventions will provide clues for possible measures to improve human health and may also give useful prognostic information. In this study, we demonstrated the following key findings: (1) prolongevity interventions generally tightened the systems-level regulation of biological processes at both transcriptional and post-transcriptional layers in mice; (2) fatty acid metabolism emerged as a common process shifted by multiple prolongevity interventions; (3) the systems-level regulation of biological processes was associated with both chronological and biological ages in humans.

By leveraging mouse omics datasets and systems-level approaches, we demonstrated that prolongevity interventions modified biological processes and metabolic reactions at the systems level (Fig. 2–5). In particular, DIRAC analyses revealed that the tightening of module regulation was a general signature of the prolongevity interventions within the measured proteomic and transcriptomic spaces (Fig. 2a, 3f, 5a). Interestingly, a previous study using DIRAC revealed the general loosening of module regulation in more malignant phenotypes and later stages of cancer progression^40^. Given that cancer resistance and longevity share commonality in mechanisms such as DNA repair and telomere maintenance^55,59^, aging may be promoted, in part, by loss of tight regulation for pertinent modules, and its tightness maintenance may be a key longevity strategy. Furthermore, we identified at least 12 proteomic modules (Fig. 2b, Supplementary Data 1), 1,829 transcriptomic modules (Supplementary Fig. 3a, Supplementary Data 5), and 1,822 reactions (Fig. 4a, Supplementary Data 3) affected by any of the prolongevity interventions. These modules and reactions included the biological processes highly related to “aging hallmarks”^55^ and “pillars of aging”^1^, such as amino acid regulation, fatty acid regulation, TCA cycle, stress response, and inflammation, consistent with the proposed roles of these processes in aging^6–9,55^. Therefore, our current study showed the power of systems-level approaches to explore and test hypotheses about the control of aging and longevity in mammals, and provided a translational implication that potential prolongevity interventions may be identified and evaluated based on their regulatory effects on these systems.

Fatty acid β-oxidation is the catabolic process of fatty acid breakdown for energy production, with mitochondria and peroxisomes being the major involved organelles^60^. We demonstrated that fatty acid β-oxidation was tightened in both proteins and transcripts consistently across mechanistically distinct prolongevity interventions (Fig. 2d, 5c). We also observed that the system transporting proteins into peroxisomes was tightened in proteins consistently across the interventions (Supplementary Fig. 1c), and implied the possibility that some aspects of mitochondrial functions were affected by 17aE2 and Rapa (Fig. 3e). Moreover, we showed that reactions involved in fatty acid synthesis and oxidation were concordantly shifted across MR, GHRKO, and SnellDW (Fig. 4d, e, Supplementary Fig. 2a, b). All these findings support the conclusion that fatty acid β-oxidation was directed towards tight control in a whole cellular system for longevity. At the same time, this systems-level control of fatty acid β-oxidation was observed quite possibly through different mechanisms by each intervention. For example, the tightening pattern in acetyl-CoA synthesis, which is essentially connected to fatty acid β-oxidation, was similar between 17aE2 and Rapa, but different from ACA (Supplementary Fig. 1b); 17aE2 and Rapa, but not ACA, similarly modulated expression patterns of mitochondrial proteins (Fig. 3b); the prolongevity interventions other than MR, GHRKO, and SnellDW did not show the (significant) flux changes in fatty acid β-oxidation (Fig. 4d). Hence, we hypothesize that different prolongevity interventions lead to a similar rerouting of energy metabolism through fatty acid metabolism, albeit through different mechanisms. Although the findings from DIRAC, WGCNA and GEM do not indicate the functional direction for cells (e.g., tight regulation can be either augmentation or attenuation of a pathway), there are multiple reports about fatty acid oxidation in aging and longevity; AMPK, an essential kinase of the nutrient-sensing signaling pathways in longevity, inhibits fatty acid synthesis and promotes fatty acid oxidation via inhibition of acetyl-CoA carboxylase 1 (ACC1) and ACC2^6,61^; CR increases fatty acid synthesis in adipose tissue but results in enhancing whole-body oxidation^62^; ketogenic diet specifically upregulates the genes involved in fatty acid oxidation in liver^15^; overexpression of fatty acid-binding protein (FABP) or dodecenoyl-CoA delta-isomerase (DCI), corresponding to the acceleration of fatty acid β-oxidation, increased lifespan in *D. melanogaster*^63^. Therefore, tight regulation promoting fatty acid β-oxidation could be a common signature among prolongevity strategies. On the other hand, the prominence of the nutrient-sensing or energy-producing process from liver-derived datasets might be unsurprising because the liver is a major metabolic organ. However, we also observed that prolongevity interventions tightened the modules less often associated with liver and metabolism, such as *RIG-I signaling* (GO:0039529; Supplementary Fig. 1e, f, 3f) and *CD40 signaling* (GO:0023035; Supplementary Data 1). In the mid-life human female brain, metabolic and immune systems are shifted by chronological age: glucose metabolism and fatty acid β-oxidation are attenuated and enhanced, respectively, and chronic low-grade innate and adaptive immune responses are enhanced^64^. Hence, the interrelationship between fatty acid metabolism and innate/adaptive inflammation is an interesting area for future investigations.

Aging accompanies progressive loss of homeostasis. This intuition can be qualitatively assessed by “allosteric load” (also known as “physiological dysregulation”), and this measure increases with chronological age^65–67^. Hence, we anticipated that the tightness of module regulation would monotonically decrease with chronological age. However, we observed the “U-shaped” trajectory of the RCI median: the tight module regulation decreased as a function of chronological age up to mid-life, and increased from mid-life onwards (Fig. 6a, Supplementary Fig. 5c). Our consecutive analyses (Fig. 6b, f, Supplementary Fig. 5d, 6c) implied that the tight patterns of module regulation were representatively characterized by a young healthy state (Y-subQ1) and an old unhealthy state (O-subQ5). Namely, one of the interpretations is that the U-shaped trajectory was deduced from the higher variations in health state among middle-aged individuals. These findings suggest that the binary interpretation of “dysregulation” is insufficient for understanding molecular and physiological processes in aging. As a limitation, we cannot deny potential effects of survivorship bias on the state transition observed in older ages, which may have diminished the monotonic reduction pattern. A further limitation is that we cannot deduce exactly which mechanisms are responsible for regulating the systems entropy that we measure (i.e., rank conservation); these mechanisms are predominantly under autonomous intracellular controls such as biochemical and transcriptional regulations but could be affected by behavioral/neurological or external/environmental controls, which may have generated the “U-shaped” pattern. Nevertheless, we have previously reported that healthy individuals have an increasingly divergent gut microbiome compositional state with age^68^. A bacterial microbiome in *D. melanogaster* is necessary for age-dependent changing patterns in metabolism and immune response^69^. Hence, there may be a critical link between the aging patterns of module regulation and gut microbiome, especially in metabolomic space^70^, which lead to the tight module regulation in older ages.

There are several limitations to this study. In DIRAC and GEM analyses, we pooled female and male samples due to a small sample size. Hence, it is highly possible that we failed to identify sex-dependent changes, especially related to the known sex-dependent effects of ACA, 17aE2, and Protandim on lifespan extension^19–21^. Because this study successfully validated the utility of systems-level approaches and because sex dimorphism in aging and longevity remains not fully elucidated^71^, we plan to address this point as a continued study by leveraging the upcoming datasets that are generated from experiments with larger sample sizes. Additionally, there were marked differences in study design between the mouse and human datasets (Fig. 1); the former addressed the systems that were changed by prolongevity interventions under young adults, while the latter addressed the systems that were observed across age in an adult population. To directly link the findings from mice to humans, one could theoretically compare the DIRAC metrics between short-lived and long-lived individuals or between the individuals with or without prolongevity intervention across decades, for example, although this is not so easy in practice. In addition, the regulation for biological systems may not be conserved between mouse liver and human blood. Moreover, there were few commonly examined modules between mouse liver proteomics and human blood proteomics due to the difference in the measured proteins (Supplementary Data 1, 8). Hence, our findings in mice and humans may be entirely unrelated. However, a coherent explanation may be possible to connect our mouse and human findings. Given that the median lifespans of UM-HET3 mice in our experimental facility are approximately 886 and 863 days for females and males, respectively, the mice used in this study (12 months old) completed around 42% of their potential lifespan. Assuming 80 years as a median lifespan for humans, this would correspond to roughly 34 years-old humans. Therefore, the module state tightened by the prolongevity interventions in mice (Fig. 2, 3, and 5) may be related to the young healthier state observed in humans (Y-subQ1 in Fig. 6 and Supplementary Fig. 6), in line with the clinical anticipation that appropriate interventions (e.g., prolongevity drug administration, dietary CR) can slow aging in humans, at least, at young or middle stage. Further investigations, including how prolongevity interventions affect older mice, are required to deepen our understanding of systems-level regulation.

## Methods

### Mouse liver proteomic datasets

Liver samples from mice fed with lifespan-extending drugs were collected as previously described^57^. Briefly, 12 (6 female and 6 male) genetically heterogeneous UM-HET3 mice were prepared for each sample group: control, acarbose (ACA), 17α-estradiol (17aE2), and rapamycin (Rapa). The drugs were treated via daily feeding of the Purina 5LG6 diet with ACA (1,000 mg kg^−1^), 17aE2 (14.4 mg kg^−1^), or Rapa (14 mg kg^−1^) starting at 4 months. At 12 months, the mice fasted for 18 h and were euthanized for liver sampling. Excised livers were washed in phosphate-buffered saline (PBS) and snap-frozen for proteomic analysis. All procedures followed the methods recommended by the National Institute on Aging (NIA) Interventions Testing Program (ITP)^18^. Hereinafter, this experiment is called “LC-M001”.

Liver samples from 4EGI-1-treated mice were collected as previously described^56^. Each group, control and 4EGI-1, consisted of 4 female and 4 male UM-HET3 mice aged 6 to 8 months old. Controls received an intraperitoneal injection of 15 µL dimethyl sulfoxide (DMSO) daily for 5 days, and treated mice received DMSO containing 4EGI-1 at 75 mg per kg body weight. After the last injection, the mice were fasted for 18 h prior to euthanasia. Excised livers were washed in PBS and snap-frozen for proteomic analysis. Hereinafter, this experiment is called “LC-M004”.

The frozen livers were dissected, processed with lysis and trypsin digestion, and analyzed by mass spectrometry (MS) for quantitative protein abundance. Liver sections were placed in lysis buffer (50 mM tris(hydroxymethyl)aminomethane (Tris)-HCl pH 8.0 and 5% sodium dodecyl sulfate (SDS)) and homogenized using a Precellys® 24 tissue homogenizer (Bertin Technologies SAS, Montigny-le-Bretonneux, France). For each sample, protein concentrations were determined by a bicinchoninic acid (BCA) assay. 300 µg of solubilized protein extract in 5% SDS was purified to remove SDS using Midi-S-Trap^™^ sample processing technology (ProtiFi, New York, USA), and digested with trypsin at 37 °C for 4 h. The extracted tryptic peptides were subjected to reverse phase liquid chromatography tandem mass spectrometry (LC-MS/MS), using an Easy-nLC 1000 (Thermo Fisher Scientific, Massachusetts, USA) with a 50 cm fused silica capillary (75 µm inner diameter) packed with C18 (ReproSil-Pur 1.9 µm; Dr. Maisch GMBH, Ammerbuch, Germany) heated to 45 °C. The mobile phase gradient consisted of 5–35% acetonitrile and 0.1% formic acid over 3 h for the LC-M001 samples or over 2 h for the LC-M004 samples. The LC-M001 samples were analyzed on a Q Exactive-HF mass spectrometer (Thermo Fisher Scientific) in data-dependent acquisition (DDA) mode with an MS scan mass range of 375–1375 *m/z* and a resolution of 60,000. MS/MS scans were acquired with TopN = 15 using 15,000 resolution, with an isolation width of 1.8 *m/z*, AGC set to 100,000, and 100 ms injection time. NCE was set to 27, and dynamic exclusion was set to 20 s. The LC-M004 samples were analyzed on an Orbitrap Fusion Lumos (Thermo Fisher Scientific) in DDA mode with an MS scan mass range of 375–1375 *m/z* and a resolution of 60,000. MS/MS scans were acquired with TopN = 12 using 15,000 resolution with an isolation width of 1.8 *m/z*, AGC set to 40,000, and 30 ms injection time. NCE was set to 30, and a dynamic exclusion was set to 30 s.

MS data analysis was conducted using the Trans-Proteomic Pipeline^72^. Peptide identification was performed by database searching with Comet^73^ using the mouse reference proteome UP000000589 (UniProt, downloaded on June 11, 2019) filtered to one protein sequence per gene. Peptide sequences were validated with PeptideProphet^74^ and iProphet^75^. Protein inference was performed with ProteinProphet^76^. Protein quantification was performed using the top-3 method^77,78^ on quantities obtained from the extracted ion chromatograms of the precursor signals of the identified proteotypic peptides.

### Mouse liver transcriptomic dataset

The processed dataset of mouse liver transcriptomics was kindly provided by Vadim N. Gladyshev (Harvard Medical School). Complete descriptions are found in the original paper^45^. Briefly, the original experiment was designed to investigate eight prolongevity interventions: two genetically modified models (the growth hormone receptor knockout mouse (GHRKO) and the hypopituitary Snell dwarf mouse (SnellDW)), two nutritional interventions (calorie restriction (CR) and methionine restriction (MR)), and four pharmacological interventions (ACA, 17aE2, Protandim®, and Rapa). Three 5 months-old male mice were prepared for each sample group in genetically modified models: SnellDW control (SnellWT), SnellDW, GHRKO control (GHRWT), and GHRKO. Three UM-HET3 mice were prepared for each of the 22 sex-and age-distinguished sample groups in nutritional and pharmacological interventions: 6 months-old female of control, CR, ACA, 17aE2, Protandim, and Rapa; 12 months-old female of control, CR, ACA, and Rapa; 6 months-old male of control, CR, ACA, 17aE2, Protandim, and Rapa; 12 months-old male of control, CR, ACA, and Rapa; 14 months-old male of MR control and MR. The nutritional and pharmacological interventions were treated via daily feeding of the Purina 5LG6 diet with CR (40% less than control) starting at 4 months, with MR (0.12% w/w methionine; cf. 0.86% w/w methionine in MR control) starting at 2 months, or with ACA (1,000 mg kg^−1^), 17aE2 (14.4 mg kg^−1^), Protandim (1,200 mg kg^−1^), or Rapa (42 or 14 mg kg^−1^ for 6 or 12 months-old, respectively) starting at 4 months. The liver samples were processed for paired-end RNA sequencing using NovaSeq 6000 sequencing system (Illumina, California, USA). The processed reads after the quality filtering and adapter removal were mapped to gene and counted. After filtering out genes with low number of reads, the count data of the filtered genes was passed to the relative log expression (RLE) normalization.

### Human plasma proteomic and metabolomic datasets

The original human plasma proteomic and metabolomic datasets relied on a cohort consisting of over 5,000 individuals who participated in the Arivale Scientific Wellness program (Arivale, Washington, USA). Complete descriptions are found in the previous papers^47,48,70^. Briefly, an individual was eligible for enrollment if the individual was over 18 years old, not pregnant, and a resident of any US state except New York; participants were primarily recruited from Washington, California, and Oregon. In this program, multiomic data was collected, including human genomes, longitudinal measurements of clinical lab tests, proteomics, metabolomics, gut microbiomes, and wearable devices, and health/lifestyle questionnaires. Peripheral venous blood draws for all measurements were performed by trained phlebotomists at LabCorp (Laboratory Corporation of America Holdings, North Carolina, USA) or Quest (Quest Diagnostics, New Jersey, USA) service centers. Proteomic data was generated using proximity extension assay (PEA) for plasma derived from whole blood samples with several Olink Target panels (Olink Proteomics, Uppsala, Sweden), and measurements with the Cardiovascular II, Cardiovascular III and Inflammation panels were used in the present study since the other panels were not necessarily applied to all samples. Metabolomic data was generated using ultra-high-performance liquid chromatography-tandem mass spectrometry (UHPLC-MS/MS) for plasma derived from whole blood samples by Metabolon (North Carolina, USA). This study was conducted with deidentified data of the participants who had consented to the use of their anonymized data in research. All procedures were approved by the Western Institutional Review Board (WIRB) with Institutional Review Board (IRB) (Study Number: 20170658 at Institute for Systems Biology and 1178906 at Arivale).

In this study, we selected the participants for whom the multiomic biological age^48^ and general covariates (Body Mass Index (BMI) and ancestry principal components (PCs)) had been calculated, and retrieved the baseline proteomic or metabolomic dataset (i.e., the first time point measurement for each participant). Analytes which were missing in more than 10% of participants were removed, and participants who had missing values for more than 10% of the remaining analytes were removed. Missing values were imputed with random forest using Python missingpy library (version 0.2.0). Some proteins were measured on multiple Olink panels; these values were averaged to produce one value per protein. The final preprocessed proteomic and metabolomic datasets were 263 proteins × 2,714 participants and 739 metabolites × 1,899 participants, respectively.

### Weighted Gene Coexpression Network Analysis

Weighted Gene Coexpression Network Analysis (WGCNA) was performed using R WGCNA package (version 1.69) according to the WGCNA methodlogy^42^. Analytes were initially filtered based on missing values with the default threshold setting (50%), and the remained analytes were used to generate the coexpression network. Network generation was performed using Spearman’s correlation and the signed-hybrid approach within the WGCNA package. The *β* parameter to approximate a scale-free topology was defined with 7 for the LC-M001 proteomics and 11 for the Arivale metabolomics, using the pickSoftThreshold function. Module identification was subsequently performed using the topological overlap matrix and the default hierarchical clustering approach with dynamic tree cut. Consequently, nine modules were identified for the LC-M001 proteomics (Fig. 3a) and the Arivale metabolomics (Supplementary Fig. 5b). The identified modules were summarized with “module eigengene”: the *q*-module eigengene *E*^(*q*)^ corresponds to the first PC of the expression matrix of proteins in that module. In addition, intramodular connectivity (i.e., the sum of the adjacency to the other nodes within the module) was calculated for each protein of the modules.

### Differential Rank Conservation analysis

#### Preprocessing

To apply Differential Rank Conservation (DIRAC) analysis, missingness in the mouse datasets was conservatively resolved by filtering out the analytes that were not detected in one or more samples; the final number of analytes was 2,231 proteins for DIRAC analysis of the LC-M001 proteomics, 2,112 proteins for DIRAC analysis of the LC-M001 and LC-M004 proteomics, and 11,192 transcripts for DIRAC analysis of the M001-related transcriptomics. Missingness in the human datasets was resolved by imputation during the aforementioned cohort-defining pipeline. In this study, the analyte values were normalized using “robust *Z*-score” (i.e., *Z*-score using median and median absolute deviation (MAD) instead of mean and SD, respectively) for each sample, and further normalized using robust *Z*-score for each analyte based on the median and MAD of the control group (mouse datasets) or the whole population (human datasets). In mouse datasets, samples with different conditions in sex and age but the same intervention were handled as a single sample group to calculate robust DIRAC rank consensus from small sample size, while recognizing the false negative risks for potential sex or age-dependent changes.

#### Module set preparation

For each protein in the preprocessed datasets, the Gene Ontology Biological Process (GOBP) annotations were retrieved using the European Molecular Biology Laboratory’s European Bioinformatics Institute (EMBL-EBI) QuickGO application programming interface (API) with a query of UniProt ID (January 26, 2021 for mouse datasets; June 1, 2022 for human dataset). For each gene in the preprocessed dataset, the GOBP annotations were retrieved using R org.Mm.eg.db package (version 3.12.0) with a query of the Ensembl ID. Each GOBP term defines a priori module consisting of all annotated proteins/genes in the corresponding species (i.e., backgrounds). To maintain the biological meaning of annotation, the modules were further selected if at least half of the members in the module, with a minimum of four members, were quantified in the preprocessed datasets; the final a priori module set was 164 modules for DIRAC analysis of the LC-M001 proteomics (Supplementary Data 1), 153 modules for DIRAC analysis of the LC-M001 and LC-M004 proteomics (Supplementary Data 7), and 3,747 modules for DIRAC analysis of the M001-related transcriptomics (Supplementary Data 5). Due to the small number of quantified proteins in the preprocessed Arivale proteomics, the selection criterion was relaxed to 30% of the proteins in each module but still at least four proteins; the final a priori module set was 19 modules for DIRAC analysis of the Arivale proteomics (Supplementary Data 8).

Data-driven modules were prepared by applying WGCNA to each of the LC-M001 proteomics and the Arivale metabolomics, as described above. Because missingness was differently handled between DIRAC analysis and WGCNA, each WGCNA-identified module could have the analytes that were not retained in the preprocessed datasets for DIRAC analysis (Fig. 3a, Supplementary Fig. 5b). Hence, the WGCNA modules were further selected if at least half of the members in the module, with a minimum of four members, were retained in the preprocessed datasets; the final data-driven module set was seven modules for DIRAC analysis of the LC-M001 proteomics (Supplementary Data 2) and nine modules for DIRAC analysis of the Arivale metabolomics (Supplementary Data 9).

#### DIRAC calculation

The DIRAC algorithm^40^ was reimplemented in Python (version 3.7.6 or 3.9.7). Briefly, pairwise comparisons of analyte values within a module are initially performed for each sample, generating a “ranking/ordering dataframe” which contains binary values about whether analyte_*i*_ value is larger than analyte_*j*_ value. Next, consensus of the binary values is calculated per analyte_*i*_–analyte_*j*_ pair for each sample group (called “phenotype” in the original paper) by majority vote, generating a binary “ranking/ordering template dataframe” which corresponds to the “rank” consensus in the DIRAC algorithm. Then, each analyte_*i*_–analyte_*j*_ pair in the ranking/ordering dataframe is judged whether it matches or mismatches with a consensus in the ranking/ordering template dataframe. Rank matching score (RMS) for each module against each consensus is obtained per sample by calculating a ratio of the number of matched pairs. Finally, RMSs for each module against each consensus is summarized with the arithmetic mean per sample group. When the mean of RMSs in a sample group is based on the consensus of the sample group itself, it corresponds to rank conservation index (RCI); that is, RCI is a special case of the RMS mean.

### Genome-scale metabolic model reconstruction

For each of the samples in the M001-related transcriptomics, a “context-specific” (i.e., sample-specific) metabolic network model was reconstructed from a mouse genome-scale metabolic model (GEM), iMM1865^43^, which is a knowledge-based multi-compartment model consisting of 1,865 metabolic genes, 10,612 reactions, and 5,839 metabolites. According to the gene–protein–reaction (GPR) associations, the RLE values were integrated with the generic iMM1865 for each sample using the integrative metabolic analysis tool (iMAT) algorithm^79^. Subsequently, to predict the flux values of reactions at steady state, flux variability analysis (FVA) was performed for each context-specific GEM using the COBRA toolbox (version 3.0)^80^. FVA evaluates the flux range for each reaction by optimizing all the potential flux distributions to minimize or maximize a pre-defined objective function under the solution space (i.e., under the context-specific constraints), which is known as the LP (Linear Programming) and MILP (Mixed Integer Linear Programming) problems. In this study, the biomass reaction (BIOMASS_reaction) defined in the generic iMM1865 was used as the objective function to be maximized, and FVA was performed for 90% of the optimal solution using the fastFVA function. COBRA toolbox was implemented in MATLAB (R2019a), and academic licenses of Gurobi optimizer (version 7.5) and IBM CPLEX (version 12.7.1) were used to solve LP and MILP. As a result, the flux ranges were successfully predicted for 7,930 reactions among the 10,612 reactions defined in the generic GEM, and the maximum value was representatively used as the predicted flux value in this study (Supplementary Data 3).

### Statistical analysis

Almost all processing and null hypothesis testing were performed using R (version 4.1.1) with R tidyverse (version 1.3.1), multcomp (version 1.4.19), dunn.test (version 1.3.5), and clusterProfiler (version 4.2.2)^81^ packages, while correlation tests, ordinary least squares (OLS) regression analyses, and preprocessing for them were performed using Python (version 3.7.6 or 3.9.7) with Python NumPy (version 1.18.5 or 1.21.3), pandas (version 1.0.5 or 1.3.4), SciPy (version 1.4.1 or 1.7.1) and statsmodels (version 0.11.1 or 0.13.0) libraries. *P* < 0.05 was considered statistically significant in all analyses. Group statistics (e.g., sample size, mean, SEM) and test summary (e.g., test statistic, exact *P*-value) are found in Supplementary Data 1–9.

For comparing overall RCI distributions, differences in the mean of RCIs between control and each intervention were assessed using two-sided Dunnett’s test (Fig. 2a, 3f, 5a) or repeated two-sided Student’s *t*-tests with the multiple hypothesis adjustment by the Holm–Bonferroni method (Fig. 5e). For identifying the module changed by any of the interventions, the intervention effect on RCI (i.e., the mean of RMSs under the rank consensus of own sample group) was assessed using Analysis of Variance (ANOVA; RMS ∼ intervention) for each module (Fig. 2b, 2c, 3g, Supplementary Fig. 3a, 3b) or each module and each omics (Fig. 5b), while adjusting multiple hypotheses with the Benjamini–Hochberg method. Note that GOBP modules are partly dependent on each other because the same gene/protein can be shared between GOBP terms; hence, this simple adjustment approach could inflate false negatives, and is regarded as a conservative approach. Additionally note that sex was not included in the ANOVA models since RMS and RCI themselves were calculated from the rank consensus of pooled groups, as described above. For subsequently clarifying which intervention changed (tightened or loosened) the module, the post hoc comparisons for RCI between control and each intervention were assessed using two-sided Dunnett’s test (Fig. 2b–d, 3g, 3h, 5h, Supplementary Fig. 1a, 1c, 1e, 3a–c, 3g, 3j) or repeated two-sided Student’s *t*-tests with the multiple hypothesis adjustment by the Holm–Bonferroni method (Fig. 5b–d). For examining the similarity of module regulation among interventions (Fig. 2c, 2e, 3i, Supplementary Fig. 1b, 1d, 1f, 3b, 3d), differences in the mean of RMSs between control and each intervention were assessed for each rank consensus using two-sided Dunnett’s test. Note that the sample group corresponding to the rank consensus group was excluded from these tests, because its mean of RMSs (i.e., RCI) is expected to follow different distribution from the other sample groups’ one. For identifying the module whose regulation similarity to the LC-M004 sample groups (Control-2, 4EGI-1) was different among the LC-M001 sample groups (Control-1, ACA, 17aE2, Rapa), the intervention effect on the RMS mean was assessed using ANOVA (RMS ∼ intervention) for each module and each rank consensus, while adjusting multiple hypotheses with the Benjamini–Hochberg method (i.e., a conservative approach, as described above). For subsequently clarifying which intervention (similarly or dissimilarly) changed the module (Fig. 5f, 5g, Supplementary Fig. 3e, 3f, 3h, 3i), the post hoc comparisons for the RMS mean between Control-1 and each intervention (ACA, 17aE2, Rapa) were assessed using repeated two-sided Student’s *t*-tests with the multiple hypothesis adjustment by the Holm–Bonferroni method. For examining whether the RCI median was dependent on the characteristics of each group (Supplementary Fig. 4b), two-sided statistical significance of the RCI median was assessed using a permutation test where an empirical null-hypothesis distribution of the RCI median was estimated from 20,000 DIRAC re-calculations with the shuffles of sample–group correspondence, while adjusting multiple hypotheses with the Holm–Bonferroni method. For examining overall relationship between the similarity of module regulation and stratified group, Spearman’s correlation between the mean of RMSs and the quantile order of stratified groups was assessed for each sex and each rank consensus (Fig. 6b, Supplementary Fig. 5d) or each sex, each chronological age tertile (CA3) group, and each rank consensus (Fig. 6f, Supplementary Fig. 6c), while adjusting multiple hypotheses with the Holm–Bonferroni method. For identifying the module whose regulation similarity was associated with chronological age (Fig. 6c, 6d, Supplementary Fig. 5e, 5f), RMS was regressed to chronological age with BMI and ancestry PC1–5 as covariates (RMS ∼ chronological age + log(BMI) + PC1 + PC2 + PC3 + PC4 + PC5) for each module, each sex, and each rank consensus, while adjusting multiple hypotheses with the Benjamini–Hochberg method (i.e., a conservative approach for GOBP modules, as described above). For identifying the module whose regulation similarity was associated with Δ age (Fig. 6g, Supplementary Fig. 4d, 6d, 6e), RMS was regressed to Δ age with chronological age, BMI, and ancestry PC1–5 as covariates (RMS ∼ Δ age + chronological age + log(BMI) + PC1 + PC2 + PC3 + PC4 + PC5) for each module, each sex, each CA3 group, and each rank consensus, while adjusting multiple hypotheses with the Benjamini–Hochberg method (i.e., a conservative approach for GOBP modules, as described above).

For identifying the WGCNA module changed by any of the interventions, the intervention effect on the module eigengene was assessed using ANOVA (*E*^(*q*)^ ∼ intervention + sex + intervention × sex) for each module, while adjusting multiple hypotheses with the Bonferroni method. For subsequently clarifying which intervention changed the module eigengene (Fig. 3b), the post hoc comparisons for the *E*^(*q*)^ mean between control and each intervention were assessed using two-sided Dunnett’s test. For examining the relationship between the intervention effect on each protein in the module and their respective intramodular connectivity (Fig. 3d), the main effect of intervention on each protein *k* was calculated using ANOVA (Protein_*k*_^(*q*)^ ∼ intervention + sex + intervention × sex), and then Spearman’s correlation between the calculated main effect of intervention and intramodular connectivity was assessed.

For identifying the reaction whose flux was changed by any of the interventions (Fig. 4a), the intervention effect on the flux value was assessed using Kruskal–Wallis *H*-test (flux value ∼ intervention) for each reaction, while adjusting multiple hypotheses with the Benjamini–Hochberg method. Note that reactions in GEM are partly dependent on each other because the same gene/protein/metabolite can be shared between the reactions; hence, this simple adjustment approach could inflate false negatives, and is regarded as a conservative approach. Additionally note that samples were pooled per intervention to increase the statistical power from small sample size, while recognizing the false negative risks for potential sex or age-dependent changes. For subsequently clarifying which intervention changed the reaction flux (Fig. 4b–d), the post hoc comparisons for the flux value median between control and each intervention were assessed using two-sided Dunn’s test with the multiple hypothesis adjustment by the Holm–Bonferroni method. For examining the difference between CR and MR (Supplementary Fig. 2c–f), the additional post hoc comparisons were assessed between CR and its control, between MR and its control, and between CR and MR. For examining which subsystems in GEM were shifted by ACA, MR, GHRKO, and SnellDW (Fig. 4e, Supplementary Fig. 2a, 2b) or differently shifted between CR and MR (Supplementary Fig. 2g), enrichment in the significantly changed reactions was assessed using overrepresentation test for each of the subsystems that were annotated to any of the significantly changed reactions, while adjusting multiple hypotheses with the Benjamini–Hochberg method.

## Data visualization

Almost all results were visualized using Python (version 3.7.6 or 3.9.7) with Python matplotlib (version 3.1.3 or 3.4.3), seaborn (version 0.10.1 or 0.11.2), venn (version 0.1.3) libraries, while the results of enrichment analyses were visualized using R (version 4.1.1) with R ggplot2 (version 3.3.6) and enrichplot (version 1.14.2) packages. The results were summarized as the mean with 95% confidence interval (CI) or the boxplot, as indicated in each figure legend. Note that this 95% CI of mean or median was simultaneously calculated during visualization using the seaborn barplot or boxplot API, respectively; hence, this CI is not exactly same with that used in statistical analysis but for presentation purpose only. Hierarchical clustering was simultaneously performed during visualization using seaborn clustermap API with the Ward’s linkage method for Euclidean distance. For the values used in Fig. 4a, the group mean of flux values for each reaction was centered by subtracting the group mean of the corresponding control, and then scaled by the maximum absolute value across intervention groups using MaxAbsScaler of Python scikit-learn library (version 1.0.1). In the scatterplots with regression lines, the adjusted sample RMS with the covariates was calculated as the mean ± residual using the OLS linear regression for each plot that was the same used in statistical analysis except for dropping the independent variable (i.e., chronological age in Fig. 6d, Supplementary Fig. 5f; Δ age in Supplementary Fig. 4d, 6e), and the regression line with 95% CI was simultaneously computed during visualization using the seaborn regplot API.

## Data Availability

The MS data of the LC-M001 and LC-M004 proteomics have been deposited to the ProteomeXchange Consortium via the PRIDE partner repository (PXD035255). Note that this data will be available after journal publication; until then, requests should be directed to the corresponding authors. The processed data of M001-related transcriptomics was kindly provided by Vadim N. Gladyshev (Harvard Medical School), and raw data is available on the NCBI's Gene Expression Omnibus (GEO) repository (GSE131901). The Arivale datasets can be accessed by qualified researchers for research purposes. Requests should be sent to. The de-identified data will be available to the qualified researchers on submission and approval of a research plan.

## Data availability

The MS data of the LC-M001 and LC-M004 proteomics have been deposited to the ProteomeXchange Consortium via the PRIDE partner repository (PXD035255)^82^. Note that this data will be available after journal publication; until then, requests should be directed to the corresponding authors. The processed data of M001-related transcriptomics was kindly provided by Vadim N. Gladyshev (Harvard Medical School), and raw data is available on the NCBI’s Gene Expression Omnibus (GEO) repository (GSE131901). The Arivale datasets can be accessed by qualified researchers for research purposes. Requests should be sent to. The de-identified data will be available to the qualified researchers on submission and approval of a research plan.

## Code availability

Code used in this study is freely available in GitHub (https://github.com/longevity-consortium).

## Acknowledgements

We thank Vadim N. Gladyshev (Harvard Medical School) for kindly providing the processed dataset of mouse liver transcriptomics; Eric S. Orwoll (Oregon Health and Science University) and Gilbert S. Omenn (University of Michigan) for providing comments to the manuscript; Jennifer Dougherty and Mary Brunkow (Institute for Systems Biology) for their coordination efforts in the National Institute on Aging (NIA) Longevity Consortium; all Arivale participants who consented to using their deidentified data for research purposes. This work was funded by the National Institutes of Health (NIH) grant U19AG023122 awarded by NIA (to S.R.C., N.J.S., R.A.M., R.L.M., and N.R.), the M.J. Murdock Charitable Trust (Reference No. 2014096:MNL:11/20/2014, awarded to N.D.P. and L.H.), and a generous gift from Carole Ellison (to K.W. and T.W.). K.W. was supported by The Uehara Memorial Foundation (Overseas Postdoctoral Fellowships).

## Author Contribution

K.W., T.W., P.B., M.R., R.A.M., R.L.M., and N.R. conceptualized the study. K.W., T.W., P.B., M.R., and N.R. participated in the study design. G.G.G. and R.A.M. performed mouse experiments. M.R.H., M.K.M., D.H.B., M.M., S.R.M., K.M.C., C.K., U.K., and R.L.M. contributed to the generation of the mouse proteomics. J.C.L. and A.T.M. managed the logistics of data collection and integration for the human datasets. K.W., T.W., and P.B. performed data analysis and figure generation. J.W., J.L., C.L., J.C.R., G.G., S.R.C., N.J.S., N.D.P., and L.H. assisted in result interpretation. K.W., T.W., P.B., M.R., and N.R. were the primary authors of the paper, with contributions from all other authors. All authors read and approved the final manuscript.

