## Supplementary Information for "Systems-level patterns in biological processes are changed under prolongevity interventions and across biological age"

**This PDF file includes:**

Supplementary Figures 1 to 6
Supplementary Data legends

**Other Supplementary Information files in this study include the followings:**

Supplementary Data 1 to 9

### Supplementary Figures

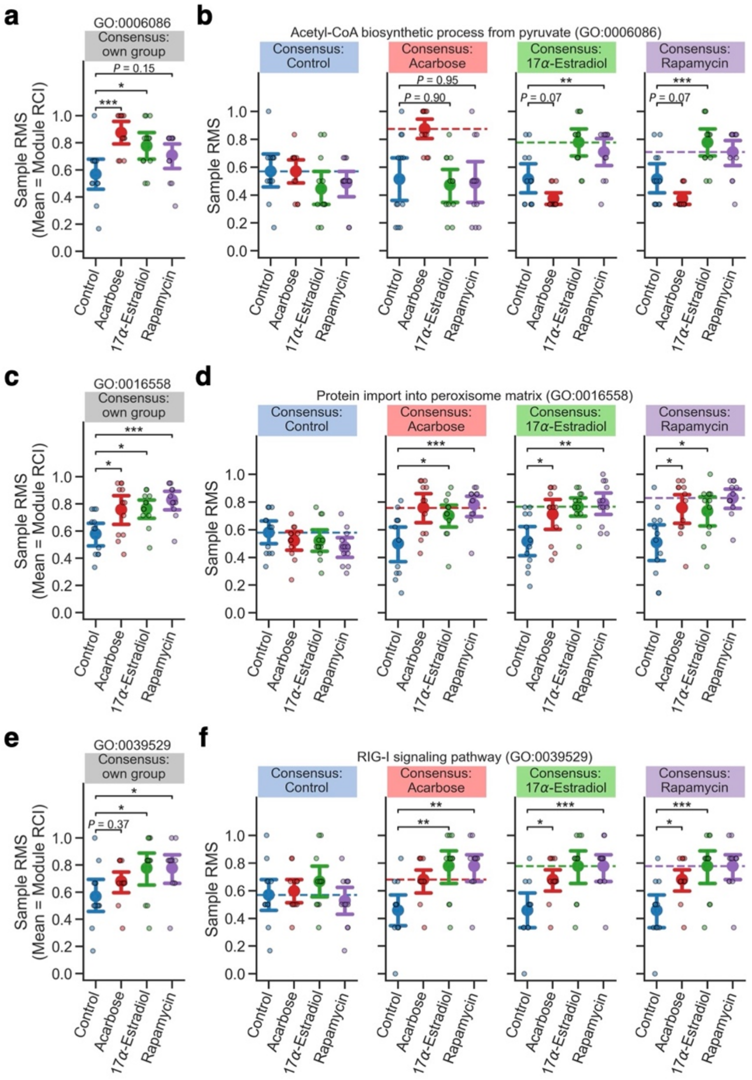


**Supplementary Figure 1. Examples of the tightened proteomic modules by prolongevity drugs.**

Shown are results from Differential Rank Conservation (DIRAC) analysis of the LC-M001 liver proteomics using Gene Ontology Biological Process (GOBP)-defined modules, corresponding to Fig. 2. **a**–**f** Sample rank matching score (RMS) distributions for examples of the tightly but differently tightened modules between intervention groups (**a**, **b**; GO:0006086, acetyl-CoA biosynthetic process from pyruvate) or the similarly tightened modules (**e**–**f**; GO:0016558, protein import into peroxisome matrix; GO:0039529, RIG-I signaling pathway). Dashed line in **b**, **d**, and **f** indicates the mean of RMSs for the sample group corresponding to the rank consensus (i.e., rank conservation index, RCI). Data: the mean (dot) with 95% confidence interval (CI, bar); $n = 12$ mice. *$P < 0.05$, **$P < 0.01$, ***$P < 0.001$ by two-sided Dunnett's test.

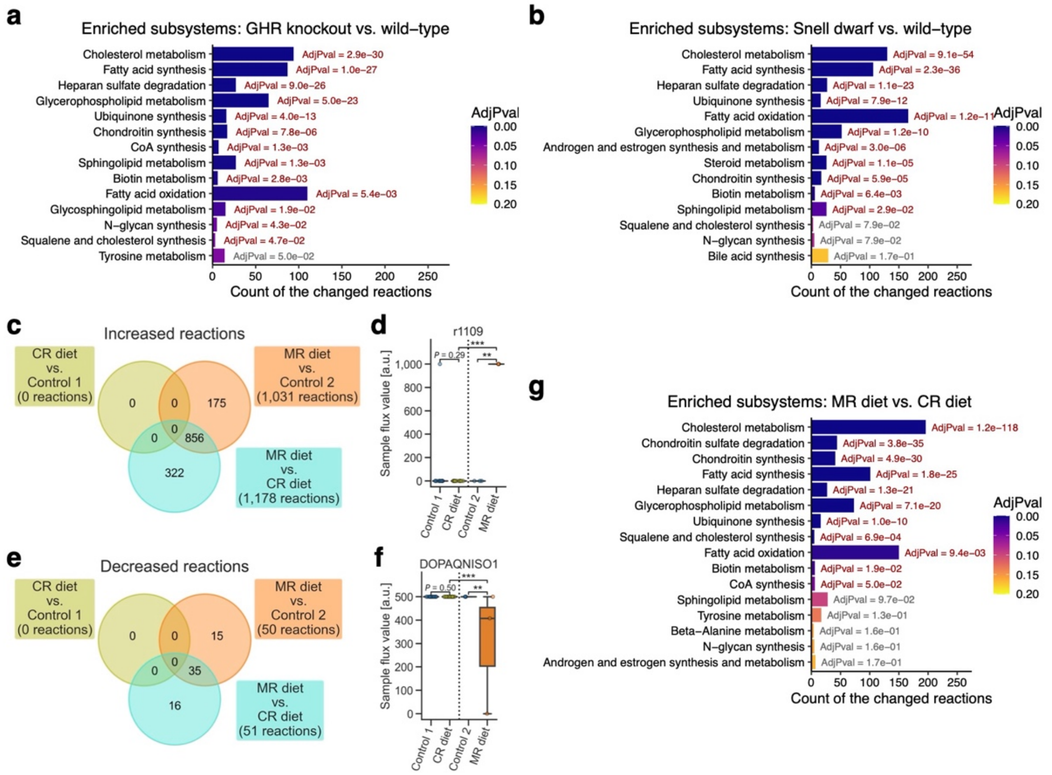


**Supplementary Figure 2. Shifted subsystems and the differences between CR and MR dietary interventions.**

Shown are results from an integrated analysis of the M001-related liver transcriptomics with mouse genome-scale metabolic model (GEM), corresponding to Fig. 4. GHR: growth hormone receptor; CR: calorie restriction; MR: methionine restriction; Control 1: control for CR diet; Control 2: control for MR diet. **a**, **b** Shifted subsystems by GHR knockout (**a**) or Snell dwarf (**b**). Significance of the shifted subsystems was assessed using enrichment analysis on the significantly changed reactions (conservatively false discovery rate (FDR)-adjusted $P < 0.05$) while adjusting the multiple hypotheses with the Benjamini–Hochberg method. Only the subsystems that exhibited nominal $P < 0.05$ are presented. AdjPval: FDR-adjusted $P$-value from the enrichment analysis. **c**–**f** Reactions exhibiting different flux values between CR and MR dietary interventions. **c**, **e** Venn diagram of the reactions that exhibited "conservatively" FDR-adjusted $P < 0.05$ (see Methods) for the main effect of intervention on each flux median by Kruskal–Wallis $H$-test and that exhibited significantly different flux median in contrast group from baseline group (i.e., "changed" reaction; $P < 0.05$ by post hoc two-sided Dunn's test with the Holm–Bonferroni adjustment). The latter group of the comparison label corresponds to the baseline group (e.g., CR diet was set as the baseline group in MR diet vs. CR diet). **d**, **f** Flux value distributions for examples of the significantly changed reactions by MR diet. Data: the 25$^{th}$ percentile ($Q_1$, box bottom), median (center line), and the 75$^{th}$ percentile ($Q_3$, box top); whiskers span [max($x_{min}$, $Q_1 - 1.5 \times$ IQR), min($x_{max}$, $Q_3 + 1.5 \times$ IQR)], where $x_{min}$ and $x_{max}$ are the minimum and maximum, respectively, in the observed values and IQR = $Q_3 - Q_1$; $n$ = 12 (Control 1, CR diet), 3 (Control 2, MR diet) mice. **$P < 0.01$, ***$P < 0.001$ by two-sided Dunn's test with the Holm–Bonferroni adjustment. **g** Different subsystems between CR and MR dietary interventions. Significance assessment of the different subsystems and figure representation are the same with **a** and **b**.

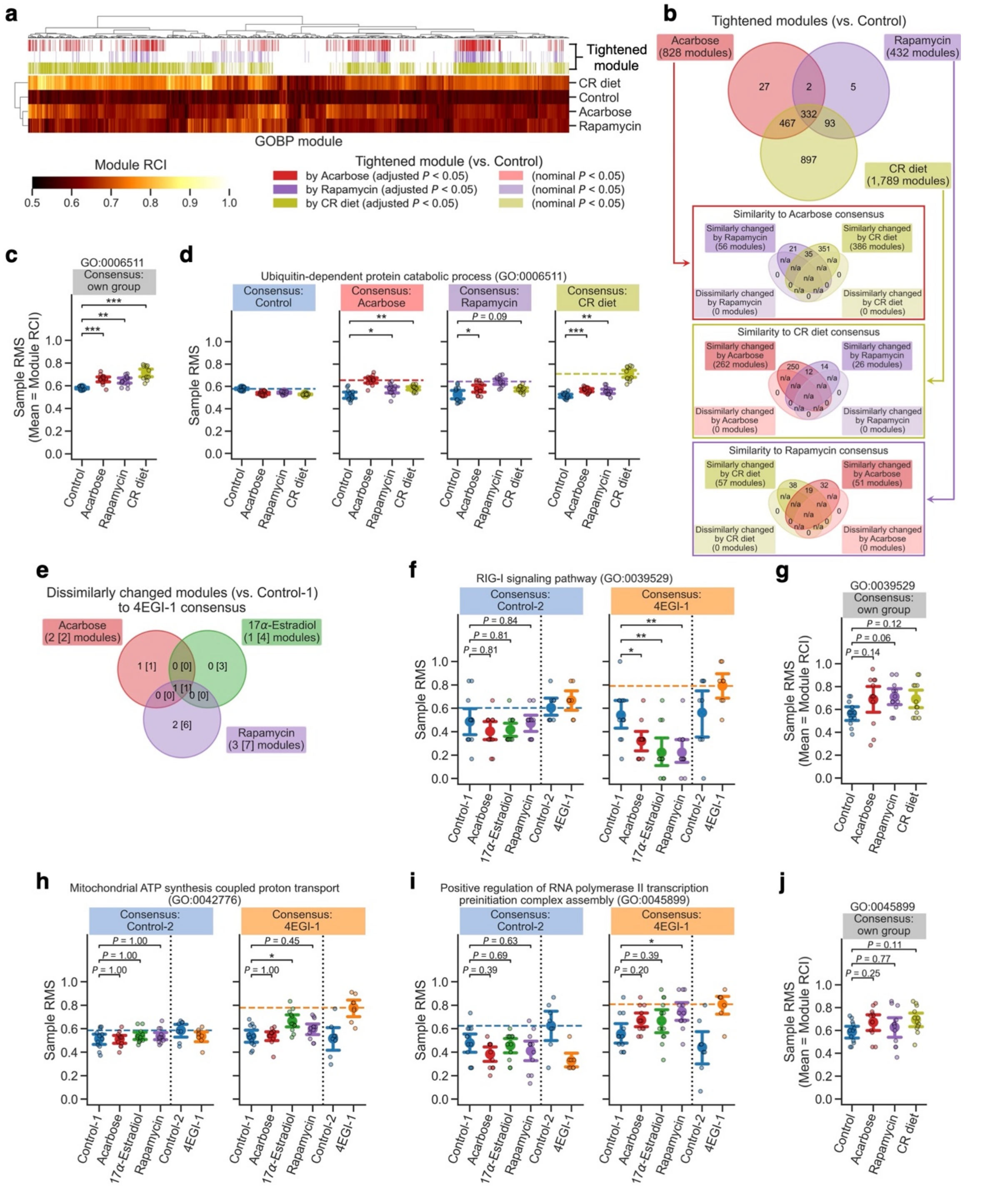


**Supplementary Figure 3. DIRAC results of the M001-related liver transcriptomics and examples of the tightened modules potentially through cap-independent translation.**

**a–d**, **g**, **j** Differential Rank Conservation (DIRAC) analysis of the M001-related liver transcriptomics using Gene Ontology Biological Process (GOBP)-defined modules, corresponding to Fig. 5a. **a** Overall distribution of module rank conservation index (RCI). Top color columns highlight the modules that exhibited nominal or "conservatively" false discovery rate (FDR)-adjusted $P < 0.05$ (see Methods) for the main effect of intervention on each module RCI by Analysis of Variance (ANOVA) and that exhibited significantly higher RCI in intervention group than control group (i.e., "tightened" module; $P < 0.05$ by post hoc two-sided Dunnett's test). **b** Venn diagrams of the significantly tightened modules by each intervention (conservatively FDR-adjusted $P < 0.05$). For the tightened modules in each intervention group, sub-venn diagram indicates the modules for which the other intervention groups exhibited significantly higher or lower mean of rank matching scores (RMSs) under the rank consensus than control group (i.e., "similarly" or "dissimilarly" changed module to the consensus group, respectively; $P < 0.05$ by two-sided Dunnett's test). n/a: logically not available. **c**, **d** Sample RMS distributions for an example of the tightened modules (GO:0006511, ubiquitin-dependent protein catabolic process). Dashed line in **d** indicates the mean of RMSs for the sample group corresponding to the rank consensus (i.e., RCI). Data: the mean (dot) with 95% confidence interval (CI, bar); $n = 12$ mice. *$P < 0.05$, **$P < 0.01$, ***$P < 0.001$ by two-sided Dunnett's test. **e**, **f**, **h**, **i** DIRAC analysis of the LC-M001 and LC-M004 liver proteomics using GOBP-defined modules, corresponding to Fig. 5e–g. Control 1: control for Acarbose, 17α-Estradiol, and Rapamycin; Control 2: control for 4EGI-1. **e** Venn diagram of the modules that exhibited nominal or "conservatively" FDR-adjusted $P < 0.05$ (see Methods) for the main effect of intervention on each module mean of RMSs under 4EGI-1 rank consensus by ANOVA and that exhibited significantly lower mean of RMSs in intervention group than control group (i.e., "dissimilarly" changed module to the 4EGI-1 group; $P < 0.05$ by post hoc two-sided Student's *t*-tests with the Holm–Bonferroni adjustment). The number in square brackets corresponds to the dissimilarly changed modules (nominal *P*-value < 0.05). **f**–**j** Sample RMS distributions for examples of the dissimilarly tightened modules (**f**, **g**; GO:0039529, RIG-I signaling pathway) or the similarly tightened modules (**h**–**j**; GO:0042776, mitochondrial ATP synthesis-coupled protein transport; GO:0045899, positive regulation of RNA polymerase II transcription preinitiation complex assembly) in proteins (**f**, **h**, **i**) or transcripts (**g**, **j**). Dashed line in **f**, **h**, and **i** indicates the mean of RMSs for the sample group corresponding to the rank consensus (i.e., RCI). Data: the mean (dot) with 95% CI (bar); $n = 8$ (Control-2, 4EGI-1), 12 (the others) mice. *$P < 0.05$, **$P < 0.01$ by two-sided Student's *t*-tests with the Holm–Bonferroni adjustment (**f**, **h**, **i**) or Dunnett's test (**g**, **j**).

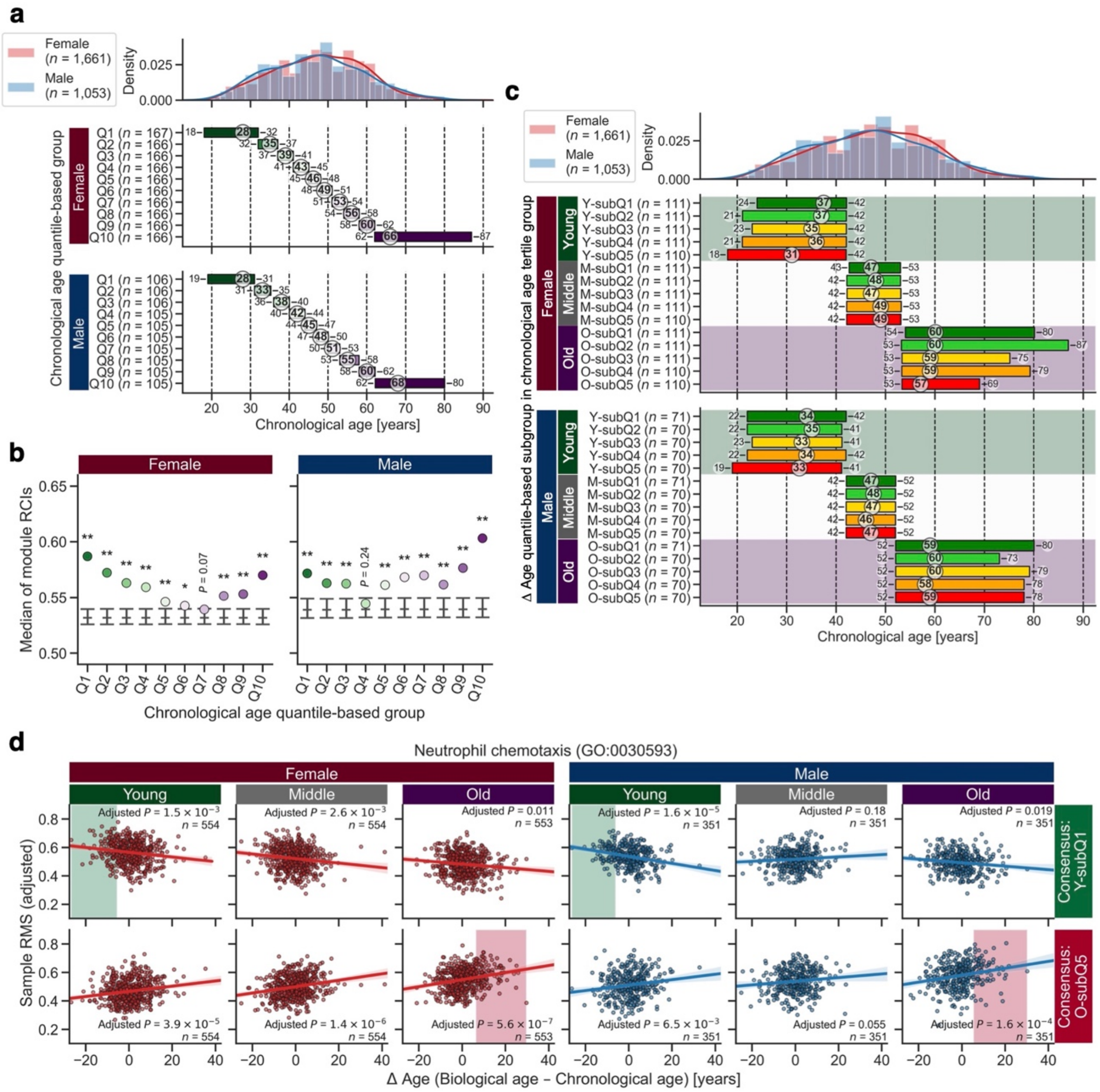


**Supplementary Figure 4. Stratified groups and DIRAC results of the Arivale plasma proteomics.**

Shown are results from Differential Rank Conservation (DIRAC) analysis of the Arivale plasma proteomics using Gene Ontology Biological Process (GOBP)-defined modules, corresponding to Fig. 6. **a** Distribution of chronological age in the chronological age-stratified groups. The numbers in the subplots indicate the minimum–median–maximum of each group. **b** Permutation test for the DIRAC patterns across chronological age-stratified groups. Each point corresponds to the median of 19 module rank conservation indexes (RCIs) in Fig. 6a. Gray bars indicate 95% confidence interval (CI) for the RCI median in a null-hypothesis distribution calculated from 20,000 randomizations, and their center ticks indicate the median. *$P < 0.05$, **$P < 0.01$ by the permutation test with the Holm–Bonferroni adjustment. **c** Distribution of chronological age in the Δ age-stratified groups. The numbers in the subplots indicate the minimum–median–maximum of each group. **d** An example of the significant association in Fig. 6g (GO:0030593, neutrophil chemotaxis). In each subplot, the adjusted sample rank matching score (RMS) with the covariates (i.e., mean ± residual) is plotted, and the colored line and background correspond to the ordinary least squares (OLS) linear regression line with 95% CI and the range of rank consensus group, respectively.

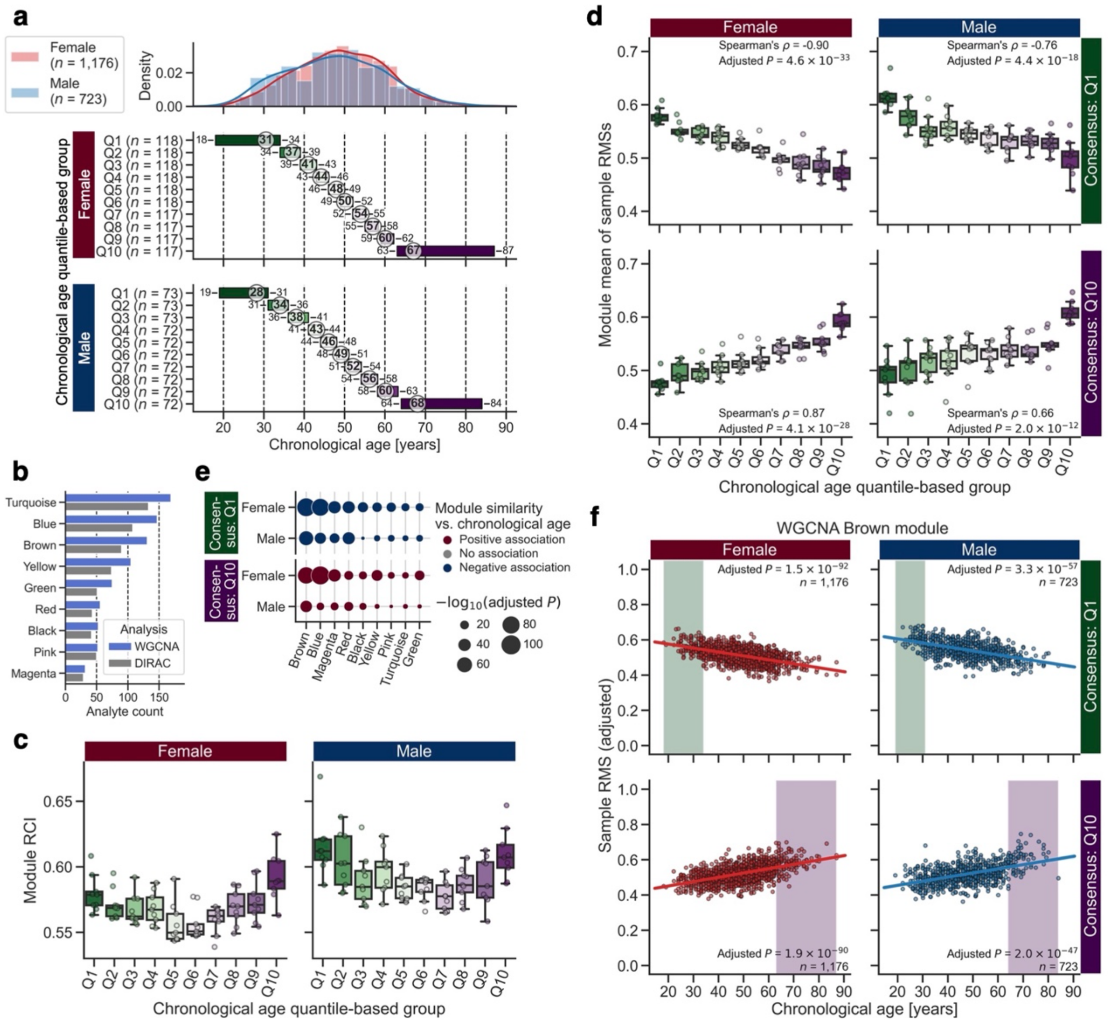


**Supplementary Figure 5. Module regulation for plasma metabolites is changed across chronological age.**

**a**–**f** Differential Rank Conservation (DIRAC) analysis of the Arivale plasma metabolomics, with the stratified groups by chronological age, using Weighted Gene Co-expression Network Analysis (WGCNA)-identified modules (see Supplementary Data 9 for complete results). **a** Distribution of chronological age in the chronological age-stratified groups. The numbers in the subplots indicate the minimum–median–maximum of each group. **b** The number of metabolites in each WGCNA-identified module. WGCNA: metabolites used in WGCNA, DIRAC: metabolites retained after the processing for DIRAC analysis. **c** Overall distribution of module rank conservation index (RCI). Data: the 25$^{th}$ percentile ($Q_1$, box bottom), median (center line), and the 75$^{th}$ percentile ($Q_3$, box top); whiskers span [max($x_{\min}$, $Q_1 - 1.5 \times$ IQR), min($x_{\max}$, $Q_3 + 1.5 \times$ IQR)], where $x_{\min}$ and $x_{\max}$ are the minimum and maximum, respectively, in the observed values and IQR = $Q_3 - Q_1$; $n = 9$ modules. **d** Overall distributions of module mean of rank matching scores (RMSs) under the rank consensus of the youngest (Q1) or oldest (Q10) group. Data: each boxplot metric is the same with **c**; $n = 9$ modules. *P*-value for Spearman's correlation was adjusted across sexes with the Holm–Bonferroni method. **e** Association between module similarity to the Q1 or Q10 group and chronological age. For each module, each rank consensus, and each sex, significance of the association was assessed using ordinary least squares (OLS) linear regression with Body Mass Index (BMI) and ancestry principal components (PCs) as covariates while adjusting multiple hypotheses with the Benjamini–Hochberg method. **f** An example of the significant association in **e** (WGCNA Brown module). In each subplot, the adjusted sample RMS with the covariates (i.e., mean ± residual) is plotted, and the colored line and background correspond to the OLS linear regression line with 95% confidence interval (CI) and the range of rank consensus group, respectively.

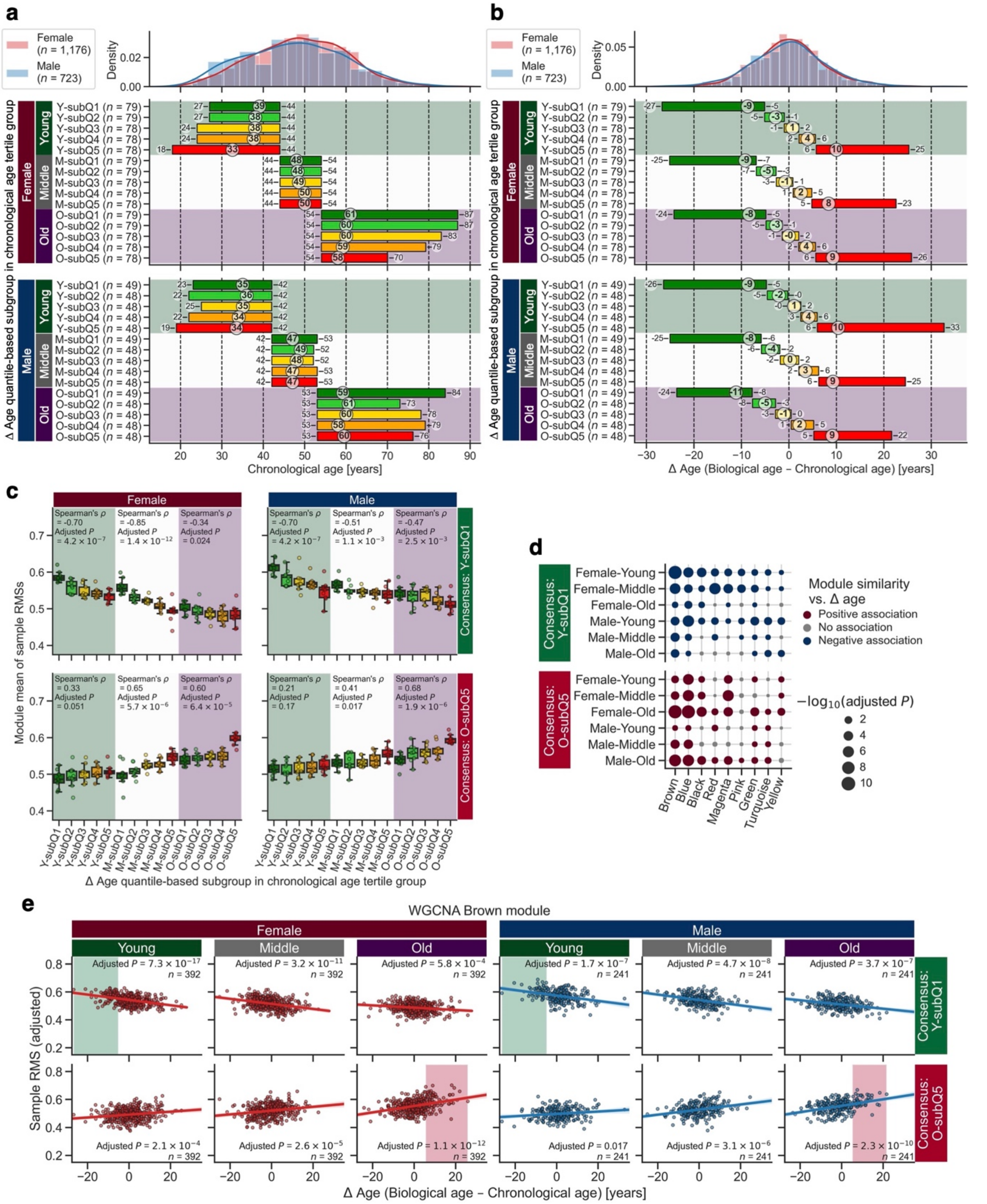


**Supplementary Figure 6. Module regulation for plasma metabolites is changed across biological age.**

**a**–**e** Differential Rank Conservation (DIRAC) analysis of the Arivale plasma metabolomics, with the stratified groups by Δ age, using Weighted Gene Co-expression Network Analysis (WGCNA)-identified modules (see Supplementary Data 9 for complete results). **a**, **b** Distribution of chronological age (**a**) or Δ age (**b**) in the Δ age-stratified groups. The numbers in the subplots indicate the minimum–

median–maximum of each group. **c** Overall distributions of module mean of rank matching scores (RMSs) under the rank consensus of the chronologically and biologically youngest (Y-subQ1) or oldest (O-subQ5) group. Data: the 25$^{th}$ percentile ($Q_1$, box bottom), median (center line), and the 75$^{th}$ percentile ($Q_3$, box top); whiskers span [max($x_{min}$, $Q_1 - 1.5 \times \text{IQR}$), min($x_{max}$, $Q_3 + 1.5 \times \text{IQR}$)], where $x_{min}$ and $x_{max}$ are the minimum and maximum, respectively, in the observed values and IQR = $Q_3 - Q_1$; $n = 9$ modules. Spearman's correlation was assessed for each of chronological age tertile (CA3) groups, and its *P*-value was adjusted across CA3 groups and sexes with the Holm–Bonferroni method. **d** Association between module similarity to the Y-subQ1 or O-subQ5 group and Δ age. For each module, each rank consensus, each sex, and each CA3 group, significance of the association was assessed using ordinary least squares (OLS) linear regression with chronological age, Body Mass Index (BMI) and ancestry principal components (PCs) as covariates while adjusting multiple hypotheses with the Benjamini–Hochberg method. **e** An example of the significant association in **d** (WGCNA Brown module). In each subplot, the adjusted sample RMS with the covariates (i.e., mean ± residual) is plotted, and the colored line and background correspond to the OLS linear regression line with 95% confidence interval (CI) and the range of rank consensus group, respectively.

### Supplementary Data Legends

**Supplementary Data 1. DIRAC analysis of the LC-M001 liver proteomics using GOBP-defined modules.**

This .xlsx file contains the DIRAC results of the LC-M001 liver proteomics using GOBP-defined modules (Fig. 2, Supplementary Fig. 1), including metadata for the examined modules, group statistics (e.g., sample size, mean, SEM), and test summary (e.g., test statistic, exact *P*-value). Descriptions about each sheet and each column are included in the README sheet.

**Supplementary Data 2. DIRAC analysis of the LC-M001 liver proteomics using WGCNA-defined modules.**

This .xlsx file contains the DIRAC results of the LC-M001 liver proteomics using WGCNA-defined modules (Fig. 3f–i), including metadata for the examined modules, group statistics (e.g., sample size, mean, SEM), and test summary (e.g., test statistic, exact *P*-value). Descriptions about each sheet and each column are included in the README sheet.

**Supplementary Data 3. Comparisons of the reaction fluxes using mouse GEM with the M001-related liver transcriptomics.**

This .xlsx file contains the comparison results of the predicted flux values using mouse GEM with the M001-related liver transcriptomics (Fig. 4, Supplementary Fig. 2), including metadata for the examined reactions, group statistics (e.g., sample size, median, MAD), and test summary (e.g., test statistic, exact *P*-value). Descriptions about each sheet and each column are included in the README sheet.

**Supplementary Data 4. Enrichment analysis on the changed reactions within the context-specific GEMs.**

This .xlsx file contains the overrepresentation test results of the significantly changed reactions within the context-specific GEMs (Fig. 4, Supplementary Fig. 2), including test summary (e.g., test statistic, exact *P*-value). Descriptions about each sheet and each column are included in the README sheet.

**Supplementary Data 5. DIRAC analysis of the M001-related liver transcriptomics.**

This .xlsx file contains the DIRAC results of the M001-related liver transcriptomics using GOBP-defined modules (Fig. 5a, h, Supplementary Fig. 3a–d, g, j), including metadata for the examined modules, group statistics (e.g., sample size, mean, SEM), and test summary (e.g., test statistic, exact *P*-value). Descriptions about each sheet and each column are included in the README sheet.

**Supplementary Data 6. Comparison of DIRAC results between the LC-M001 liver proteomics and the M001-related liver transcriptomics.**

This .xlsx file contains the comparison results of DIRAC results between the LC-M001 liver proteomics and the M001-related liver transcriptomics (Fig. 5b–d), including metadata for the examined modules, group statistics (e.g., sample size, mean, SEM), and test summary (e.g., test statistic, exact *P*-value). Descriptions about each sheet and each column are included in the README sheet.

**Supplementary Data 7. DIRAC analysis of the LC-M001 and LC-M004 liver proteomics.**

This .xlsx file contains the DIRAC results of the LC-M001 and LC-M004 liver proteomics using GOBP-defined modules (Fig. 5e–g, Supplementary Fig. 3e, f, h, i), including metadata for the examined modules, group statistics (e.g., sample size, mean, SEM), and test summary (e.g., test statistic, exact *P*-value). Descriptions about each sheet and each column are included in the README sheet.

**Supplementary Data 8. DIRAC analysis of the Arivale plasma proteomics.**

This .xlsx file contains the DIRAC results of the Arivale plasma proteomics using GOBP-defined modules (Fig. 6, Supplementary Fig. 4), including demographic summary of the overall cohort and stratified groups, metadata for the examined modules, and test summary (e.g., test statistic, exact *P*-value). Descriptions about each sheet and each column are included in the README sheet.

**Supplementary Data 9. DIRAC analysis of the Arivale plasma metabolomics.**

This .xlsx file contains the DIRAC results of the Arivale plasma metabolomics using WGCNA-defined modules (Supplementary Fig. 5, 6), including demographic summary of the overall cohort and stratified groups, metadata for the examined modules, and test summary (e.g., test statistic, exact *P*-value). Descriptions about each sheet and each column are included in the README sheet.
